# EmulatRx: Empowering Clinical Trial Design with Agentic Intelligence and Real World Data

**DOI:** 10.1101/2025.04.17.25326033

**Authors:** Haoyang Li, Weishen Pan, Suraj Rajendran, Chengxi Zang, Fei Wang

## Abstract

Clinical trial design (CTD) is a time-consuming process that requires substantial domain expertise. Large-scale real-world data (RWD), such as electronic health records (EHR), encodes practice-based evidence that is of tremendous value to CTD. In recent years, many machine learning methods have been developed to extract such real-world evidence (RWE) from the RWD to inform CTD, but they still need to be communicated with the domain experts extensively in an iterative manner to be further refined and ultimately useful. In this paper, we introduce EmulatRx, an agentic framework that derives RWE for helping with CTD. Through the iterative conversation and analysis across agents with different roles, EmulatRx can autonomously refine trial protocols and finally generate a robust report containing insights that inform better CTD. We applied EmulatRx on the CTD process for both acute diseases (e.g., septic shock, acute heart failure, acute pulmonary edema, and acute kidney injury) using the MIMIC-IV data and chronic diseases (e.g., Alzheimer’s disease and Parkinson’s disease) using the INSIGHT Network across five New York City health systems. The results demonstrate EmulatRx’s capabilities in facilitating and accelerating the CTD process.

## Introduction

Randomized controlled trials (RCTs) remain the gold standard for evaluating the efficacy and safety of medical interventions. The time and costs, as well as ethical considerations of conducting a full RCT have led researchers and practitioners to seek approaches to improve the clinical trial design (CTD) process to achieve efficiency and success rate of the corresponding RCT. Real world data (RWD), such as electronic health records (EHRs) and insurance/pharmaceutical claims, contain tremendous practiced based evidence that are insightful for informing CTD. Numerous statistical and machine learning models have been developed in the past decade for extracting such real world evidence (RWE),^1–5^ among which target trial emulation (TTE)^6^ is a representative framework with the goal of emulating an RCT with RWD. By explicitly mirroring the protocol of an RCT, defining eligibility criteria, specifying treatment assignment strategies, identifying relevant start and end times for follow-up, and selecting appropriate analytic strategies, TTE aims to estimate causal treatment effects and produce results that can closely approximate what might have been derived from an RCT. Several recent research works^7–10^ have demonstrated the great potential of TTE.

Despite the promise, there are several challenges of implementing TTE. First, the elements in the target trial protocol (including eligibility criteria, treatment strategies, and outcome, etc.) need to be matched to the RWD. Usually these elements are described as natural language in the trial protocol, but the EHR data include a large portion of standardized structured information (e.g., encoded with the Observational Medical Outcomes Partnership (OMOP^1,11–13^)). Rigorous computable phenotyping process^14^ is needed to build such mappings. Second, there is information in the target trial protocol that may not exist in RWD, such as the special biomarkers associated with particular diseases. In this case, we need domain expertise to determine if the information can be dropped or effective surrogate information can be constructed from the RWD. Third, RWD are observational in nature, where the patients are not randomized. This introduces complex selection and confounding biases. Thus, appropriate covariate balancing and causal inference methods are needed to estimate the “true” treatment effects from the RWD.

In addition to replicating RCTs with RWD, TTE also offers an effective tool for extracting evidence from RWD. For example, Zang et al.^13^ used it as a hypotheses generation tool for identifying repurposable drug candidates for Alzheimer’s disease. Liu et al.^15^ leveraged it to assess the impact of different eligible criteria on real world treatment effectiveness estimation for non-small cell lung cancer trials. Rajendran et al.^16^ designed a stratified TTE study for investigating the heterogeneous response for corticosteroid treatment for sepsis patients. In these studies, TTE produced insights even though a target trial may not exist. These insights can help with the design of a real RCT, through extensive conversations with domain experts and iterative adjustments, and this process is still time-consuming.

The rapid development of multi-agent systems (MAS) provides the opportunity for building an autonomous system to derive RWE and inform CTD. MAS are computational frameworks where multiple autonomous entities, or agents, interact and collaborate to achieve a set of goals.^17^ Each agent typically has its own specialty, knowledge base, and decision-making logic. Agents are often implemented as combinations of large language models (LLMs) with access to executable tools through APIs, enabling them to carry out diverse and complex tasks. The interactions among the agents have proven effective for coordinating diverse expertise, integrating heterogeneous data sources, as well as enabling parallel and iterative problem-solving.^18–24^ The complexity of CTD made MAS a promising approach for improving its efficiency and effectiveness.

In this paper, we present EmulatRx, a multi-agent framework designed to facilitate and accelerate CTD through the automated extraction and refinement of RWE from EHRs with Reinforcement Learning from Human Feedback (RLHF^25^). The agents in EmulatRx collaborate together to finish the following procedures: (1) collect and standardize the information from existing clinical trials and literatures to construct a knowledge graph for the design of a target clinical trial; (2) generate the protocol of a target clinical trial based on the above knowledge graph; (3) map the trial related knowledge to EHR through computable phenotypes and create cohorts; (4) conduct statistical analysis to derive RWE that can inform the CTD; (5) iteratively refine the trial protocol until satisfactory. EmulatRx also leverages RLHF to iteratively improve the quality of the entire MAS with expert feedback. We further demonstrate EmulatRx’s capability through case studies on trial design for acute conditions in the ICU setting with the MIMIC-IV^26^ data set, as well as for chronic diseases using the INSIGHT Network.^27^

## EmulatRx Architecture

### Ethics Statement

This study was approved by the Institutional Review Board of Weill Cornell Medicine with protocol number 21-07023759. All EHR data used in this study were fully deidentified; therefore, ethics approval and informed consent were not required.

### Overview

EmulatRx is a modular multi-agent framework designed to support efficient, automated, and expert-aligned CTD. It comprises five specialized agents, i.e., Supervisor, Trialist, Informatician, Clinician, and Statistician, each powered by LLMs and equipped with distinct domain-specific capabilities (Figure 1a). These agents collaborate through structured conversations to perform key steps in the trial design workflow, including protocol specification, data extraction, covariate selection, statistical modeling, and interpretation. The architecture not only includes a core sequential pipeline, i.e., Supervisor → Trialist → Informatician → Clinician → Statistician → Supervisor (as shown in Figure 1c), which reflects the natural progression from initial trial specification to analysis and reporting, but also extends beyond this traditional sequential execution by supporting dynamic interactions among agents. For instance, the Informatician can consult the Clinician when facing data sparsity or missing covariates, prompting iterative refinements in eligibility criteria or variable/outcome substitutions. This flexible agent communication strategy allows the system to adapt to appropriately accommodate the challenges in RWD. Note that EmulatRx does not require manual specification of the workflow, allowing agents to dynamically respond to one another’s outputs and adapt to new constraints or evolving objectives. For instance, if the Informatician identifies high levels of missingness for a key variable or detects poor covariate balance, the system can autonomously initiate a feedback loop with the Clinician to assess alternative variable definitions or biomedically appropriate surrogates. To ensure control and reproducibility, EmulatRx acts not as an unstructured chatbot but as a structured system built on LangGraph, which enforces a strict Graph-Based Control Flow. In this context, we define an agent as a modular unit combining a role-specific LLM reasoning engine, a persistent memory state, and a set of executable tools. Agents operate as distinct nodes within a predefined graph, and transitions (handoffs) between them act as directed edges governed by explicit logic. All conversation history and data transformations are stored in a centralized, serializable state object, allowing the complete progress of tasks to be inspected and reproduced (e.g., verifying exactly why the control transitioned from the Informatician to the Clinician). Furthermore, to mitigate the stochasticity of LLMs, the framework implements an LLM Response Cache and enforces fixed random seeds for all downstream statistical tools, ensuring that experimental results can be identically replicated provided the cache is preserved. In addition, EmulatRx integrates tool-augmented reasoning capabilities of each agent (Figure 1b). A clinical trial knowledge was built from clinical trial registries on clinicaltrial.gov to help the Trialist identify relevant protocols; the RAG module empowers the Clinician to ground its decisions in biomedical literature by performing semantic searches over large corpora such as PubMed; and the Trial Simulator provides the Statistician with access to statistical and machine learning libraries for confounder adjustment, outcome analysis, and treatment effect estimation. These tools are invoked automatically within the agent workflow to convert natural language insights into executable code, structured queries, and interpretable analytics. Beyond these core utilities, EmulatRx supports advanced reasoning functions (Figure 1d), including knowledge grounding from literature to improve factual accuracy, RLHF to align agent outputs with expert preferences, eligibility criteria (EC) optimization using Shapley-based attribution methods to quantify the influence of inclusion rules on outcomes, and subgroup analyses to uncover heterogeneous treatment effects that may be masked in the aggregate population. These capabilities are modular yet synergistic, enabling EmulatRx to continuously refine trial protocols with both methodologically robust and clinical interpretability.

**Figure 1.**
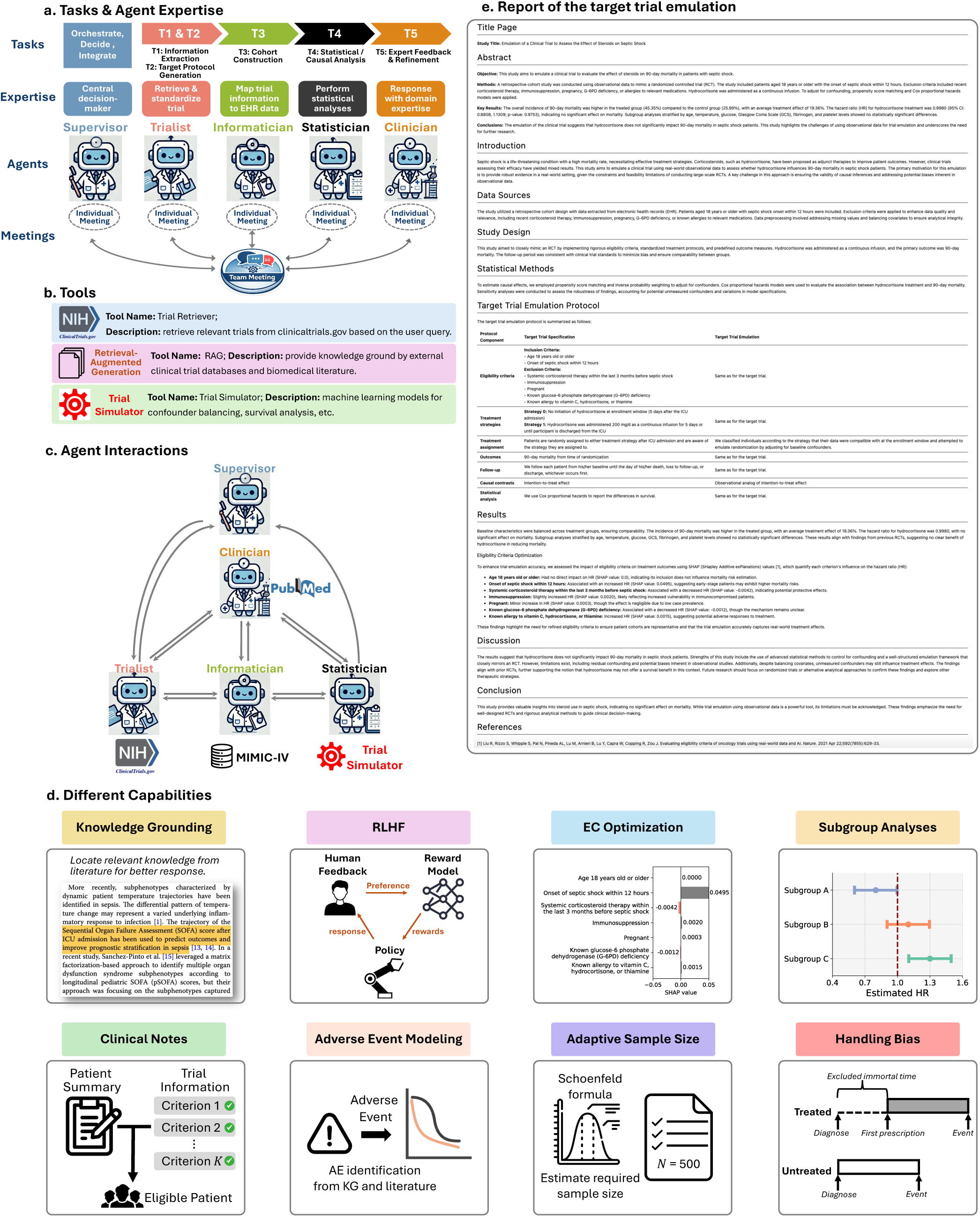
(a) Tasks and agent expertise in EmulatRx. The core tasks and their corresponding agents are: (T1) trial information extraction and standardization (Trialist), (T2) target trial protocol generation (Trialist), (T3) mapping trial specifications to EHR data and cohort construction (Informatician), (T4) statistical analysis and causal effect estimation (Statistician), and (T5) iterative refinement based on expert feedback (Clinician). The Supervisor serves as a central decision-maker that orchestrates the workflow, coordinates agent interactions, integrates intermediate results, and determines whether the process should iterate or terminate. (b) Tools used in EmulatRx. Tools include a trial retriever for querying ClinicalTrials.gov, a retrieval-augmented generation (RAG) module for grounding clinical reasoning in external literature, and a trial simulator for confounder adjustment and causal effect estimation. (c) Agent interactions. The Supervisor coordinates structured interactions among the Trialist, Informatician, Statistician, and Clinician, enabling flexible interactions and feedback loops. (d) Several Capabilities supported in EmulatRx. EmulatRx now supports knowledge grounding, RLHF, eligibility-criteria optimization, subgroup analyses, adverse event modeling, adaptive sample size estimation, and bias handling. (e) Generated final report, which mainly includes the trial emulation protocol and key results of the septic shock case.

The final output of this multi-agent system is a comprehensive trial design report (Figure 1e), which synthesizes contributions from all agents into a unified document. This report includes standardized sections, such as abstract, introduction, methods, protocol specifications, results, and discussion, and is enriched with protocol tables, statistical summaries, and visualizations (e.g., hazard ratios with confidence intervals, covariate balance diagnostics). By automating the generation of such high-quality outputs, EmulatRx not only accelerates the design cycle but also ensures transparency and reproducibility. Together, these components reflect EmulatRx’s capacity to transform the traditionally manual, expert-driven process of CTD into an efficient, intelligent, and collaborative workflow.

### Agents

Each agent in EmulatRx is designed to perform a specific set of tasks as summarized below, contributing to the overall efficiency and accuracy of the process to inform CTD.

- *<u>Supervisor</u>*: This agent acts as the central decision-maker, synthesizing inputs from the human user and determining the start or end of the workflow.
- *<u>Trialist</u>*: This agent retrieves and standardizes trial information from ClinicalTrials.gov and PubMed and then derives protocols of clinical trials to be simulated.
- *<u>Informatician</u>*: The Informatician bridges trial information with RWD, generating dataset ready for analysis. This agent matches trial eligibility criteria to EHR data, performs quality assurance, and finally constructs datasets for statistical analysis.
- *<u>Clinician</u>*: The Clinician provides domain expertise by analyzing literature and answering clinical questions. This agent is responsible to identify related covariates and outcomes, validate trial design and interpret statistical analysis results.
- *<u>Statistician</u>*: The Statistician conducts trial emulations and statistical analyses. This agent selects statistical methods, conducts outcome analyses, and generates results.

The core tasks in EmulatRx could be summarized as follows: (T1) trial information extraction and standardization, (T2) target trial protocol generation, (T3) mapping trial specifications to EHR data and cohort construction, (T4) statistical analysis and causal effect estimation, and (T5) iterative refinement based on expert feedback. These tasks are implemented by the Trialist (T1–T2), Informatician (T3), Statistician (T4), and Clinician (T5), while the Supervisor coordinates their execution and makes key decisions.

## EmulatRx for Clinical Trial Design

### Query and Summarize Clinical Trial Information

Given the user’s interest, it is critical to search for and summarize relevant information from historical trials, which is the main job of the trialist agent. To facilitate the management, retrieval, and reuse of existing clinical trials, we constructed a trial knowledge graph that contains the complete information of clinical trials.

Figure 2 illustrates the schema of the trial knowledge graph. The knowledge graph comprises nodes representing clinical trials, key trial components, and associated clinical concepts. Each trial node will correspond to a specific clinical trial and include primary metadata as node attributes, such as start and completion dates, baseline and follow-up periods, sample size, and trial phase. Component nodes will capture essential elements of clinical trials, including treatments, eligibility criteria, outcome measures (i.e., trial endpoints), and commonly used adverse events. Unlike basic trial attributes, these components are semantically richer and are represented using clinical concepts and logical, mathematical, or temporal operations. We apply design patterns from established clinical trial ontologies to represent the logical and temporal relations in the trial components.^28,29^ Each component node will include attributes such as its associated design pattern and related clinical concepts.

**Figure 2.**
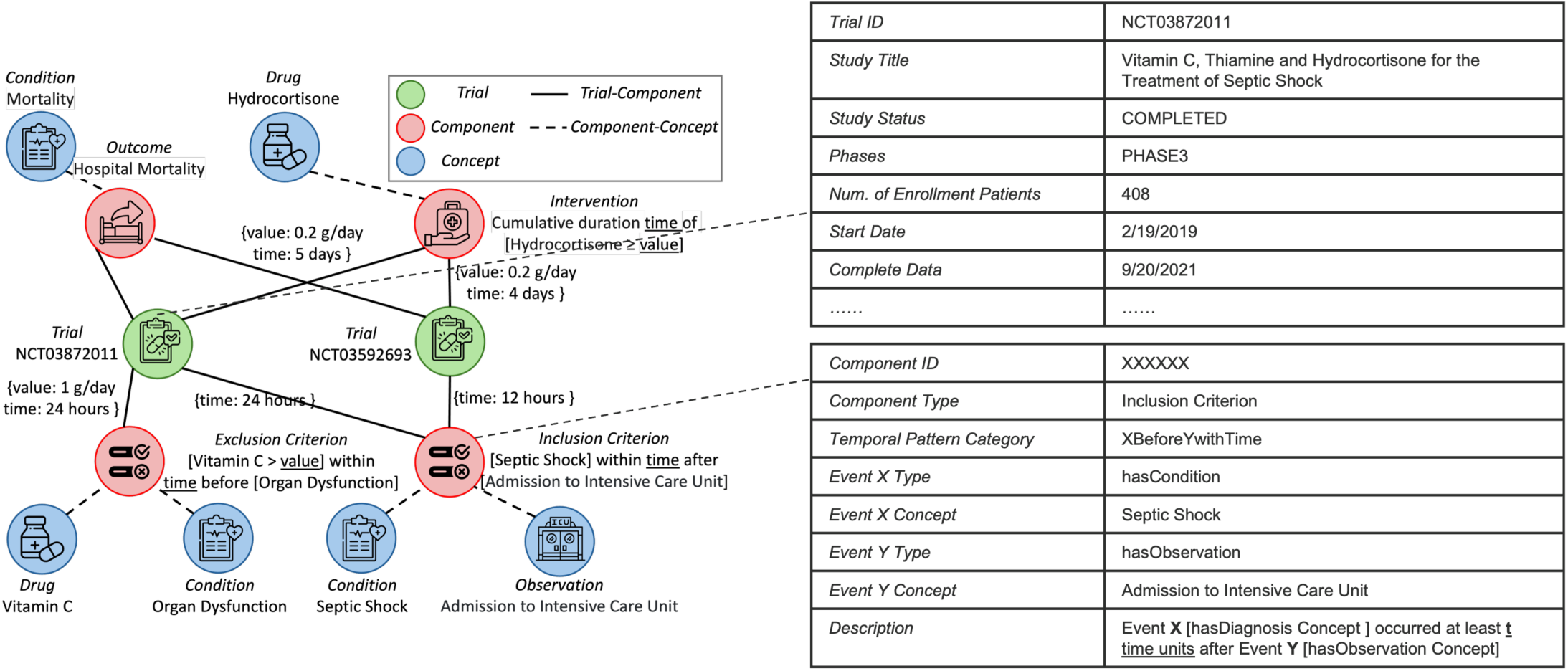
The Clinical Trial Knowledge Graph.

We constructed the knowledge graph by extracting, annotating, and standardizing key information such as eligibility criteria, treatment strategies, and outcomes. The pipeline is shown in Figure 3. First, we used an LLM concept extraction prompt, designed and refined based on the Criteria2Query3.0^30^ framework, to extract key components of the relevant trials, including eligibility criteria, treatment, and outcomes. Relevant clinical concepts are identified and annotated into domains such as Demographics, Condition, Device, Procedure, Drug, Measurement, Observation, and Visit, and their associated values along with the temporal information are extracted as well. We added additional instructions to the prompt for handling the components with multiple concepts (e.g. ““Allergy to vitamin C, hydrocortisone, or thiamine””) or omitted concepts (e.g. “Patients < 18 years” with concept age omitted in the text). The original prompt is from Criteria2Query3.0^30^ and our modified version is provided in Supplementary Table 1.

**Figure 3.**
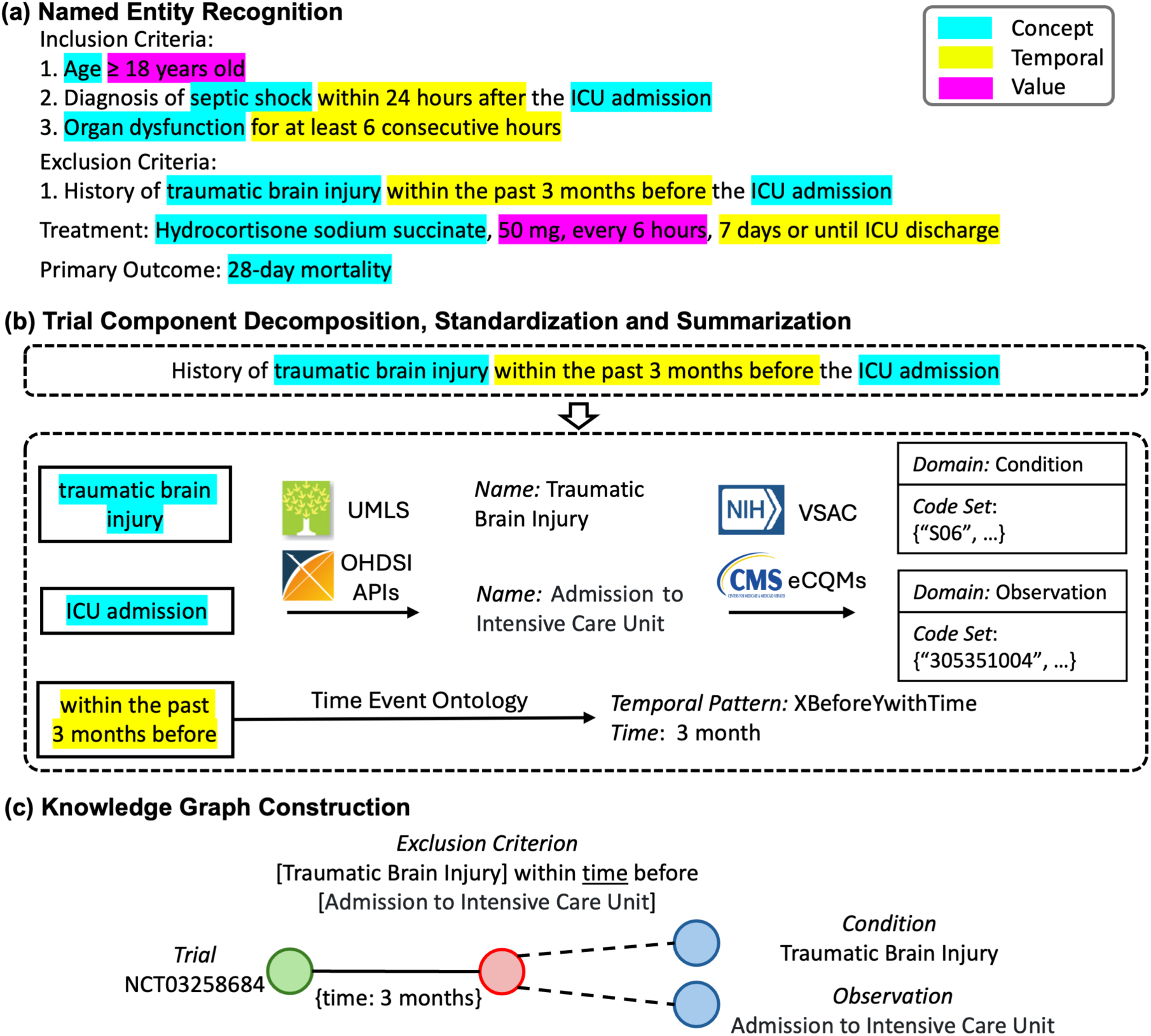
The pipeline to build the knowledge graph.

Since identical concepts might be expressed differently across trials (e.g., “ICU” vs. “Intensive Care Unit”), the Unified Medical Language System (UMLS)^31^ dictionary and Observational Health Data Sciences and Informatics (OHDSI^32^) APIs are used to standardize concepts. Then, each component is matched with design patterns defined in the existing clinical trial ontologies.^28,29^ The mathematical and temporal operators (e.g., “less than” from “less than 24 hours”), as well as the related number and units, are identified by the temporal and value normalization modules from the Criteria2Query3.0 framework^30^. All the concepts involved in the target trials are mapped to their concept ids via the standardized vocabulary in OMOP common data model. Specifically, the concepts were represented based on the standard of OMOP CDM: condition (ICD-9/ICD-10), drug (RxNorm), measurement (LOINC), procedure (SNOMED CT). This provided the necessary information for the informatician to generate the SQL queries for retrieving relevant information from the EHR data.

Leveraging the trial knowledge graph constructed through the above procedures, the Trialist agent can efficiently retrieve all relevant trials for a given study question. It supports diverse queries with complex conditions (e.g., “Retrieve all sepsis clinical trials that target ‘hydrocortisone’ as the intervention but exclude pregnant women, with a sample size greater than 100”). For the identified trials, the agent can also extract standardized representations of key information, including eligibility criteria, treatment strategies, and outcomes, from the knowledge graph. Concretely, these queries are executed against a Neo4j-backed graph database using declarative graph pattern matching, where trials, trial components (eligibility criteria, treatment strategies, and outcomes), and related concepts are represented as nodes and relation semantics are encoded as explicit edges. This Neo4j-based implementation enables efficient traversal and composition of complex query constraints across both structured trial metadata and normalized eligibility criteria, while maintaining transparency and reproducibility of the retrieval logic. When multiple trials meet the query conditions, the Trialist summarizes the results by aggregating the frequency of each unique criterion, treatment strategy, and outcome across the retrieved trials.

### Build Dataframe for Analysis

Once the trial related information has been retrieved and standardized, the Informatician agent constructs a high-quality analytical dataframe that operationalizes the trial specifications using real-world EHR data. The Informatician translates these trial related information, including eligibility criteria, treatment assignments, and outcome measures, into executable SQL queries, inspired by Criteria2Query3.0.^30^ Each criterion is rendered as a Common Table Expression (CTE),^33^ allowing for modular and sequential cohort construction. Simple eligibility rules—such as “Age ≥ 18 years”—are directly translated into SQL filters, while more complex conditions, such as time-sensitive interventions or compound dosage rules, are handled through nested logic and temporal joins.^34,30,35,36^ This structured querying ensures that the resulting cohort faithfully represents the target population defined by the trialist agent.

Beyond leveraging structured EHR data, the Informatician agent incorporates NLP techniques to analyze unstructured clinical notes, thereby enriching the patient eligibility assessment process. Structured data, while essential, may not always capture the complete clinical narrative or nuanced details pertinent to specific eligibility criteria. Clinical notes, on the other hand, contain a wealth of information regarding patient history, conditions, and observations that are often not codified.^37,38^ By processing these notes, the Informatician can identify additional eligible patients who might be missed by queries limited to structured fields alone.

The methodology for NLP-based patient matching involves a multi-stage pipeline designed to accurately ascertain eligibility from free-text notes. First, to optimize the process, an LLM (e.g., Gemma 3, Phi-4) generates a set of relevant medical keywords, synonyms, and abbreviations derived directly from the trial’s inclusion and exclusion criteria. These keywords are then used to perform an initial pre-filtering of patient notes, identifying those most likely to contain pertinent information. For these candidate notes, a more rigorous check is conducted. Both the criteria and the note sentences are text-cleaned (e.g., normalization, abbreviation expansion) and then processed using sentence embedding models (e.g., S-PubMedBert^39^) to identify the most relevant text snippets in the note for each criterion via semantic search. These note snippets (as premise) and the corresponding criterion (as hypothesis) are then fed into a biomedical natural language inference (NLI) model. The NLI model determines if the snippet entails, contradicts, or is neutral towards the criterion, with specific confidence thresholds guiding the decision of whether a criterion is met. Finally, patients who pass this NLI-based assessment for all criteria undergo a concluding verification step. A general-purpose LLM reviews the complete set of eligibility criteria alongside the full patient note to provide a final determination of eligibility, ensuring a holistic assessment before a patient is included in the emulated trial cohort.

To collect necessary covariates for downstream analysis, the Informatician collaborates with the Clinician agent, who provides advice on clinically relevant variables based on the disease context and trial objectives. These typically include demographic features, laboratory values, vital signs, diagnosis and medications. The selected covariates are then mapped to OMOP-compliant fields and integrated into the dataframe.

With the cohort and covariates defined, the Informatician can extract data from EHR warehouse to build a comprehensive, analytics-ready dataframe. Treatment data includes drug administration records, dosage schedules, and timing; outcome data focuses on time-to-event endpoints such as 28-day mortality; and covariate data spans the pre-intervention period, capturing relevant clinical measurements. All variables are aligned temporally and semantically with the trial protocol.

Data quality assurance is a critical part of this pipeline. The Informatician performs rigorous checks for completeness, logical consistency, and clinical plausibility.^40–42^ For example, outcome durations are validated to ensure non-negativity, and missingness is assessed across key variables. In cases of high missing rates, the agent may consult the Clinician to identify validated surrogate markers such as base excess. Outliers are addressed adaptively through imputation or exclusion strategies,^43,44^ depending on clinical context. The finalized output is a clean, structured dataframe containing patient-level rows with identifiers, eligibility flags, treatment indicators, outcome metrics, and baseline covariates, fully aligned with the elements needed for a trial protocol and ready for causal inference and statistical analysis.

The Informatician agent sends only the database schema and trial protocol, never patient data, to the model to generate SQL queries. These queries are then executed locally behind the institution’s firewall. This ensures that even when using proprietary cloud-based models like GPT-4o, patient privacy is preserved.

### Statistical Analysis

The Statistician agent in EmulatRx is responsible for translating the study protocol into a reproducible analytic workflow, applying appropriate statistical analysis, especially causal inference techniques, and summarizing the results. This agent’s methodology comprises five main components: (1) selecting balancing and modeling methods, (2) performing covariate balancing, (3) conducting survival analyses, and (4) generating a final report of the findings. These components are executed by a single Statistician agent operating through a multi-turn internal loop, where the agent sequentially plans the analysis, invokes the necessary tools, and retains memory of previous steps to self-correct if validation checks fail.

First, the Statistician selects the best covariate balancing strategy and outcome analysis method. In EmulatRx, the agent evaluates across multiple options for balancing including Propensity Score Matching (PSM),^45–47^ Inverse Probability of Treatment Weighting (IPTW),^48–50^ or no balancing. The Statistician bases this selection on factors such as sample size, the distribution of covariates, and the research objective of estimating causal effects, which are determined by analyzing the input from the Informatician. If the dataset is moderately sized and sufficiently rich in covariates, PSM is often chosen and preferred for its intuitive design and interpretability.^51^ To mitigate potential immortal time bias—a common issue in observational studies where treatment groups may appear to have better outcomes simply because patients had to survive long enough to receive the treatment—the Statistician agent first employs an extended clone-censor-weight methodology. This involves creating “always-treated” and “never-treated” clones for each subject, artificially censoring the never-treated clone at the observed time of treatment initiation, and then applying inverse probability of censoring weights (IPCW) to account for this informative censoring before proceeding to covariate balancing and outcome estimation.^52,53^ For each outcome analysis step, the Statistician can select from Cox Proportional Hazards,^54–56^ Kaplan-Meier estimation,^57^ parametric survival models, random survival forests (RSFs),^58^ or doubly robust methods.^59^ The final choice depends on checks of proportional hazards assumptions, the nature of the endpoints, and the need to balance interpretability (e.g., hazard ratio estimates) with predictive performance (e.g., random survival forests for high-dimensional data).

Beyond the main analytical workflow of building trial protocols, balancing covariates, and conducting survival analyses, the Statistician agent in EmulatRx can deepen the investigation of treatment effects and refine the design of the emulated trial. Specifically, the Statistician can perform subgroup analysis and EC optimization to explore treatment heterogeneity and systematically evaluate the influence of different conditions on overall results.

Following an initial survival analysis (for instance, via Cox Proportional Hazards), the Statistician agent may detect that the estimated treatment effect is not statistically significant in the overall sample. In such a scenario, the Statistician can perform subgroup analyses, wherein it prompts the Clinician to propose a covariate and threshold for splitting the cohort into two clinically relevant subgroups (e.g., patients with a severity marker below or above a certain level). The Statistician then reruns the survival model for each subgroup, comparing the hazard ratios and confidence intervals separately. This helps identify any sub-populations where the treatment might be more (or less) effective—an important consideration in critical care settings, where interventions can yield heterogeneous responses. If no subgroup exhibits a significant effect or if multiple subgroup splits fail to uncover meaningful patterns, the process stops to avoid excessive data-driven exploration.

Another critical component of the Statistician agent’s functionality is adverse event (AE) reporting, which supports safety analysis alongside treatment efficacy estimation. EmulatRx implements AE reporting with two key steps: first, defining relevant adverse events, and second, modeling these events using appropriate statistical methods. In the first stage, adverse event definitions are obtained from two sources. When the information is available in the clinical trial knowledge graph, the Trialist agent retrieves a list of common adverse events historically associated with the disease or intervention under investigation. These events are derived from previously registered and annotated with standard vocabularies to ensure compatibility with EHR data. In parallel, when adverse event details are reported in the literature associated with the target trial, the Clinician agent uses its retrieval-augmented generation capability to extract these events from the literature. This typically includes adverse reactions listed in safety endpoints or result summaries. By synthesizing trial-specific and domain-level knowledge, EmulatRx ensures that relevant and interpretable adverse event definitions are available for downstream analysis.

With the definitions of these AEs, the Statistician models the identified adverse events using methods aligned with their temporal and clinical relevance. When adverse events are considered standalone safety endpoints, they are modeled using time-to-event survival analysis with standard Cox proportional hazards models. The agent selects covariates and balancing strategies like those used for the primary outcome, ensuring comparability and consistency across analyses. The generated results include subdistribution hazard ratios, confidence intervals, and cumulative incidence functions.

Trial emulation also often requires iterative adjustments to eligibility criteria to balance sample size, comparability with the original trial, and clinical relevance. To systematically evaluate which inclusion or exclusion conditions exert the greatest impact on the estimated hazard ratio, the Statistician implements Trial Pathfinder.^60^ First, the agent enumerates different combinations (or “subsets”) of eligibility rules, computing a hazard ratio for each subset. Next, it treats the presence or absence of each criterion within these subsets as a contribution game, where Shapley values quantify how adding a particular criterion changes the measured treatment effect. If a criterion consistently shifts the hazard ratio toward or away from a significant result, it receives a larger absolute Shapley value, indicating a strong influence on outcomes. This is then relayed back to the Clinician or Supervisor, who can decide whether to retain, modify, or remove certain criteria considering both statistical impact and clinical imperatives. By systematically exploring all—or a carefully selected range of—criterion combinations, the Statistician helps ensure that trial emulation decisions are informed by quantitative evidence as well as domain expertise.

In addition to estimating treatment effects and refining eligibility criteria, the Statistician agent in EmulatRx is equipped with adaptive sample size calculation functionality to support prospective trial planning. This capability is particularly useful in early-stage design or feasibility assessment, where determining an adequate sample size is critical for ensuring sufficient power to detect a clinically meaningful effect. EmulatRx implements an automated procedure grounded in the Schoenfeld formula for CoxPH model, which estimates the required number of events and derives the total sample size based on specified hazard ratios, Type I error (α), and power (1–β). The procedure begins by examining the dataset assembled by the Informatician agent. Key characteristics are extracted, including treatment group proportions, baseline event rate in the control group, median survival time, censoring rate, and average follow-up duration. These inputs are used to estimate the cumulative event rate in the study population. EmulatRx then computes the necessary number of events using the Schoenfeld formula, where the expected hazard ratio and variability between treatment arms are incorporated through the HR and group proportions. The required sample size is subsequently derived by dividing the event count by the cumulative event rate, yielding a data-driven estimate that accounts for real-world censoring and follow-up dynamics. This adaptive procedure is fully automated within the Statistician’s workflow and can be triggered either explicitly by the other agents or implicitly when trial feasibility is in question for the Statistician. The output is the final sample size estimate. This functionality allows human experts to adjust assumptions or validate model expectations if applicable. By incorporating adaptive sample size calculation, EmulatRx extends its utility from retrospective trial emulation to prospective trial planning, offering one solution for both design validation and hypothesis generation in real-world clinical research.

Finally, the Statistician synthesizes all findings into a cohesive report. This write-up includes baseline summaries of the matched or weighted populations, biases that were adjusted for, adjusted hazard ratios (or other metrics such as risk differences, depending on the selected model), confidence intervals, and p-values. Where relevant, the Statistician compares the emulated trial estimates to published results from the original randomized trial, highlighting potential sources of discrepancy such as sample size limitations, subtle differences in inclusion criteria (due to the limitations of EHRs), or unresolved confounding. This report is then sent to the Clinician agent by the Supervisor for further review, ensuring that any remaining clinical or methodological concerns are addressed before finalizing conclusions on the treatment’s effectiveness.

### Request Clinician Suggestions

The Clinician agent in EmulatRx is designed to incorporate domain-specific medical expertise into the trial emulation workflow, ensuring that decisions about eligibility criteria, covariates, and study design remain clinically valid. Its methodology is structured around three core functions: retrieving and synthesizing medical literature, communicating clinical insights in a structured format, and collaborating iteratively with other agents to refine trial design and analysis.

The Clinician agent operates through two primary tasks: (1) reviewing reports generated by the Statistician agent and either recommending modifications or approving the analysis, and (2) providing evidence-based recommendations to other agents at various stages of the trial emulation process. These tasks are facilitated by a retrieval-augmented generation (RAG)^61^ approach to gather relevant medical literature. Upon receiving a query—such as a request to confirm clinical plausibility or identify alternative covariates—the agent performs semantic searches over a biomedical knowledge base.

Currently, this knowledge base comprises full-text PDFs or extracted abstracts stored in a FAISS index.^62^ By leveraging embeddings generated by a sentence-transformer, the Clinician agent identifies the most pertinent sections of the literature, such as published guidelines on sepsis management or prior studies on corticosteroids, and synthesizes the retrieved passages into concise, evidence-based responses.

To ensure interoperability with other agents, the Clinician agent delivers its responses in standardized, machine-readable formats. For instance, when specifying eligibility criteria, it produces annotations with tags such as <CONDITION>, <DRUG>, or <MEASUREMENT> to ensure consistent handling of clinical concepts. If recommending a relaxation of the time window for septic shock diagnosis, the agent might return an output explicitly identifying the condition domain (<CONDITION>septic shock</CONDITION>) and the temporal modifier (<TEMPORAL>within 48 hours after</TEMPORAL>). This structured output requirement functions as a guardrail against hallucination and selection bias. By forcing the Clinician agent to output standardized annotations (e.g., <CONDITION>septic shock</CONDITION>, <TEMPORAL>within 48 hours</TEMPORAL>) rather than free text, the system prevents ambiguous or fabricated concepts from reaching the data extraction phase. If the Clinician generates a condition that the Informatician cannot map to a valid OMOP concept ID, the workflow is designed to halt or automatically trigger a clarification request back to the Clinician. Additionally, the Supervisor and Informatician agents act as critics in a validation loop. If a proposed criterion results in extreme data sparsity or zero matching patients, the Informatician flags this anomaly, prompting a feedback loop where the Clinician must verify the threshold or propose a biomedically appropriate surrogate.

The Clinician agent collaborates iteratively with other agents throughout the trial emulation process. For example, when the dataset assembled by the Informatician is too small or when covariate balance is deemed insufficient by the Statistician, the Clinician proposes clinically acceptable modifications, such as relaxing exclusion criteria or substituting missing measurements with clinically equivalent variables. Similarly, for the task of report reviewing, if the Statistician identifies unexpected findings, the Clinician reviews medical literature to determine whether the results align with established knowledge, propose additional adjustments, or investigate potential explanations for the discrepancies. If the discrepancies are justifiable, the clinician adds this to the final report.

### Optimization

RLHF^63^ is deployed in EmulatRx to iteratively improve the quality of the entire MAS with expert feedback. Its workflow includes three steps: (1) human feedback collection, (2) preference modeling, and (3) policy optimization with PPO^64^ and DPO.^65^

<u>Human Feedback Collection</u>. The outputs generated by each agent are first reviewed by human experts. Then the experts provide either: *<u>Ratings</u>* (e.g., 1–5) based on task-specific criteria such as correctness, clinical validity, or alignment with research goals; or *<u>Rankings</u>*, in which they compare multiple candidate outputs and indicate preference order.

<u>Preference Modeling</u>. Within the dataset aggregated from the human feedback, rating data is used to train reward models that predict scalar-valued quality scores *r*(*x*) for outputs *x* and ranking data is converted into pairwise preferences (*x*^+^, *x*^-^), indicating that output *x*^+^ is preferred over *x*^-^ for a given task.

<u>Policy Optimization with PPO and DPO</u>. EmulatRx fine-tunes the policies of its LLMs using the following methods: (1) Proximal Policy Optimization (PPO^64^): PPO is employed when expert feedback provides scalar ratings, especially in tasks with well-defined correctness signals (e.g., SQL validity). Then the policy *Π*_0_(*x*) is optimized to maximize the expected reward:

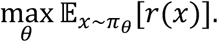

To ensure stability and prevent policy collapse, PPO uses a clipped surrogate objective:

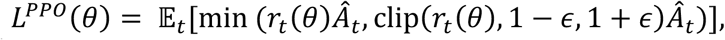

where 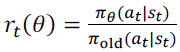 is the importance sampling ratio, 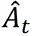 is the advantage estimate, often approximated as *r*(*x*_)_) − *b* (*b* is a baseline). (2) Direct Preference Optimization (DPO^65^): DPO is employed when feedback is provided in the form of pairwise comparisons. Rather than estimating a separate reward model, DPO directly optimizes the policy using the relative likelihoods of preferred vs. non-preferred samples. The objective is to maximize:

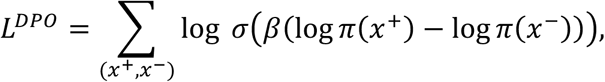

where *Π*(*x*) is the policy’s probability of output *x*, *β* is a temperature parameter controlling preference sharpness, and *σ*(⋅) is the sigmoid function.

### Meetings

In EmulatRx, meetings are designed as collaborative sessions that enable agents to share expertise, refine specific tasks, and ensure high-quality output, inspired by Swanson et al.^66^ Depending on the specific task, meetings are categorized into two types, including team meetings and individual meetings.

### Team Meetings

Team meetings in EmulatRx bring together all the agents to address complex, high-level issues requiring interdisciplinary expertise. These meetings are organized by the Supervisor, who sets the meeting agenda and synthesizes inputs. The discussions typically revolve around broad questions, such as optimizing trial eligibility criteria or selecting outcome analysis methods. For instance, consider a meeting focused on addressing high missing rates in lactate levels within the dataset. The Supervisor initiates the discussion by outlining the agenda: assessing whether surrogate variables like base excess can replace lactate levels. Each agent contributes based on their expertise. The Informatician presents data on the extent of missingness and the feasibility of implementing surrogate measures. The Clinician evaluates the clinical validity of base excess as a substitute, referencing medical literature from PubMed or other relevant sources. The Statistician weighs the statistical implications, particularly the impact on covariate balancing. The discussion unfolds over multiple rounds, with agents refining their responses based on feedback. The Supervisor consolidates the insights, approves the use of base excess, and assigns follow-up tasks to the Informatician for implementation. These meetings are essential for resolving ambiguities and achieving consensus on critical decisions. By organizing open discussions among the agents, team meetings ensure that EmulatRx leverages diverse expertise to navigate the inherent complexities of CTD.

### Individual Meetings

Individual meetings focus on task-specific activities, typically assigned to a single agent, with optional feedback from other agents or the Supervisor. These meetings allow for in-depth execution and refinement of specialized tasks, such as coding SQL queries or running outcome analysis models.

An example of an individual meeting could involve the Informatician tasked with generating a dataset based on the eligibility criteria. The agenda specifies the need to construct SQL queries that incorporate relaxed temporal conditions for septic shock diagnosis. The Informatician writes the initial SQL queries, reviews and refines the queries iteratively, and finally outputs the dataset, accompanied by a summary of modifications and explanations. These individual meetings are important for maintaining the quality of the agent’s outputs.

## Results

We evaluated EmulatRx’s performance by assessing the specialized capabilities of each agent using MIMIC-IV^26^ database and INSIGHT clinical research network^27^. We selected a diverse evaluation set of clinical trials covering various diseases, interventions, and trial phases. We curated 20 clinical trials in total, including 10 trials from MIMIC-IV focusing on acute conditions (e.g., septic shock, acute heart failure, acute pulmonary edema, and acute kidney injury) and 10 trials from INSIGHT covering chronic diseases (e.g., Alzheimer’s disease and Parkinson’s disease) with long-term follow-up and longitudinal eligibility constraints. This design enables evaluation across distinct care settings, temporal structures, and phenotype definitions. Our evaluation spanned five core dimensions: (1) clinical trial query based on the knowledge graph, (2) entity extraction and trial parsing for each trial, (3) SQL query generation by the Informatician, (4) causal inference and outcome analysis by the Statistician, and (5) clinical reasoning and recommendation quality by the Clinician. Across these tasks, we benchmarked several LLMs, including GPT-4o^67^ and three locally deployed LLMs Phi-4,^68^ DeepSeek-R1:14b^69^ (hereafter referred to as DeepSeek-R1), and Gemma-3:12b^70^ (hereafter referred to as Gemma 3), to evaluate their impact on agent performance.

### Evaluations on Trialist

For the Trialist, evaluations concentrate on (1) the query accuracy of the clinical trials based on the knowledge graph; (2) entity parsing for the trial information.

We collected 1363 clinical trials from clinicaltrial.gov, covering the diseases of septic shock, acute kidney injury, acute heart failure, and acute pulmonary edema in ICU. We evaluated the accuracy of querying relevant clinical trials with a use case as the effect of hydrocortisone in septic shock patients. We designed the following example, ranging from easy to complicated, depending on the involvement of criteria:

*Case query* 1: Retrieve all septic shock clinical trials that target “hydrocortisone” as the intervention. *Case query* 2: Retrieve all septic shock clinical trials that target “hydrocortisone” as the intervention but exclude patients with Glucose-6 phosphate dehydrogenase (G-6PD) deficiency from participating. *Case query* 3: Retrieve all septic shock clinical trials that target “hydrocortisone” as the intervention but exclude patients with platelet counts of less than 30000 per cubic millimeter from participating.

We followed existing work^71^ to employ a hybrid approach combining keyword search with manual review to identify relevant clinical trials to serve as the ground truth for each case query (details provided in Supplementary Note 1). We then compared the manually curated results with those retrieved by the automated queries with the following methods: (1) query on our trial knowledge graph; (2) query with the API of the clinicaltrial.gov; (3) query directly with the GPT-4o. For the ClinicalTrials.gov API baseline, the queries were manually constructed by a human by identifying relevant keywords from each clinical question and issuing those keywords to the ClinicalTrials.gov API. To ensure transparency and reproducibility, the exact query keywords and corresponding API calls used for each case are explicitly listed in Supplementary Note 1. The GPT-4o baseline was included as a naïve LLM-based retrieval reference to illustrate the limitations of directly using a general-purpose LLM for trial identification without external grounding. Specifically, the same clinical questions were posed to GPT-4o in a standard chat interface, and the trial identifiers returned by the model were collected and evaluated manually.

The results are presented in Table 1. With the support of the clinical trial knowledge graph, Trialist accurately identifies all relevant clinical trials across all query conditions. In contrast, directly using the ClinicalTrials.gov API can retrieve all relevant trials only when the query is limited to the target disease and treatment. Its performance declines when the query includes additional eligibility criteria, due to the lack of preprocessing and structured representation of free-text eligibility information in the original ClinicalTrials.gov database. Directly querying LLMs yields high precision, but misses many eligible clinical trials, leading to lower recall.

**Table 1.** Case query evaluation results.

|  |  | *Case query 1* | *Case query 2* | *Case query 3* |
| --- | --- | --- | --- | --- |
| # of relevant clinical trials |  | 34 | 10 | 1 |
| Knowledge Graph | # of retrieved clinical trials | 34 | 10 | 1 |
|  | Precision (%) | 100 | 100 | 100 |
|  | Recall (%) | 100 | 100 | 100 |
| API of the clinicaltrial.gov | # of retrieved clinical trials | 34 | 8 | 2 |
|  | Precision (%) | 100 | 100 | 50 |
|  | Recall (%) | 100 | 80 | 100 |
| GPT-4o | # of retrieved clinical trials | 5 | 3 | 1 |
|  | Precision (%) | 100 | 100 | 100 |
|  | Recall (%) | 14.7 | 30 | 100 |

Besides querying all the relevant trials, parsing the eligibility criteria in each trial is also essential in the following steps. For quantitative evaluation, we used the above 20 clinical trials covering both acute and chronic diseases. Eligibility criteria of the 20 selected trials were manually annotated by two biomedical informatics experts to create a gold standard, and inter-annotator agreement is measured using Cohen’s Kappa,^72^ targeting a score of ≥0.7. The evaluation was conducted at the level of knowledge graph nodes corresponding to clinical concepts parsed from trial protocols. A parsed concept node was deemed correct only when all associated attributes, including the concept name, semantic category (e.g., condition or observation), temporal qualifier, and value, exactly matched the corresponding human annotation. Metrics for entity parsing include: (1) Precision: The proportion of correctly identified concepts among all extracted concepts. (2) Recall: The proportion of correctly identified concepts out of all concepts in the gold standard. (3) F1-score: The harmonic mean of Precision and Recall.

Results are presented in Figure 4a. A total of 266 concepts were annotated across the 20 selected clinical trials. GPT-4o achieved the best performance in identifying these concepts, with a recall of 98.9% and precision of 96.7%. The high recall indicates that most of the ground-truth concepts (263 out of 266) can be correctly recognized. In comparison, the highest performance among the other models was achieved by Gemma, with a recall of 88.0% and precision of 91.8%. Notably, GPT-4o significantly outperformed other models in handling components that involve multiple concepts or omitted scopes, especially when enhanced with our prompt augmentation strategy. For example, in the criterion “Allergy to vitamin C, hydrocortisone, or thiamine,” GPT-4o accurately extracted the concepts “allergy to vitamin C,” “allergy to hydrocortisone,” and “allergy to thiamine,” while other models returned fragmented or incomplete extractions such as “allergy,” “vitamin C,” “hydrocortisone,” and “thiamine.” In another case, for the criterion “patients < 18 years,” GPT-4o correctly inferred the omitted concept “age,” which other models failed to recognize.

**Figure 4.**
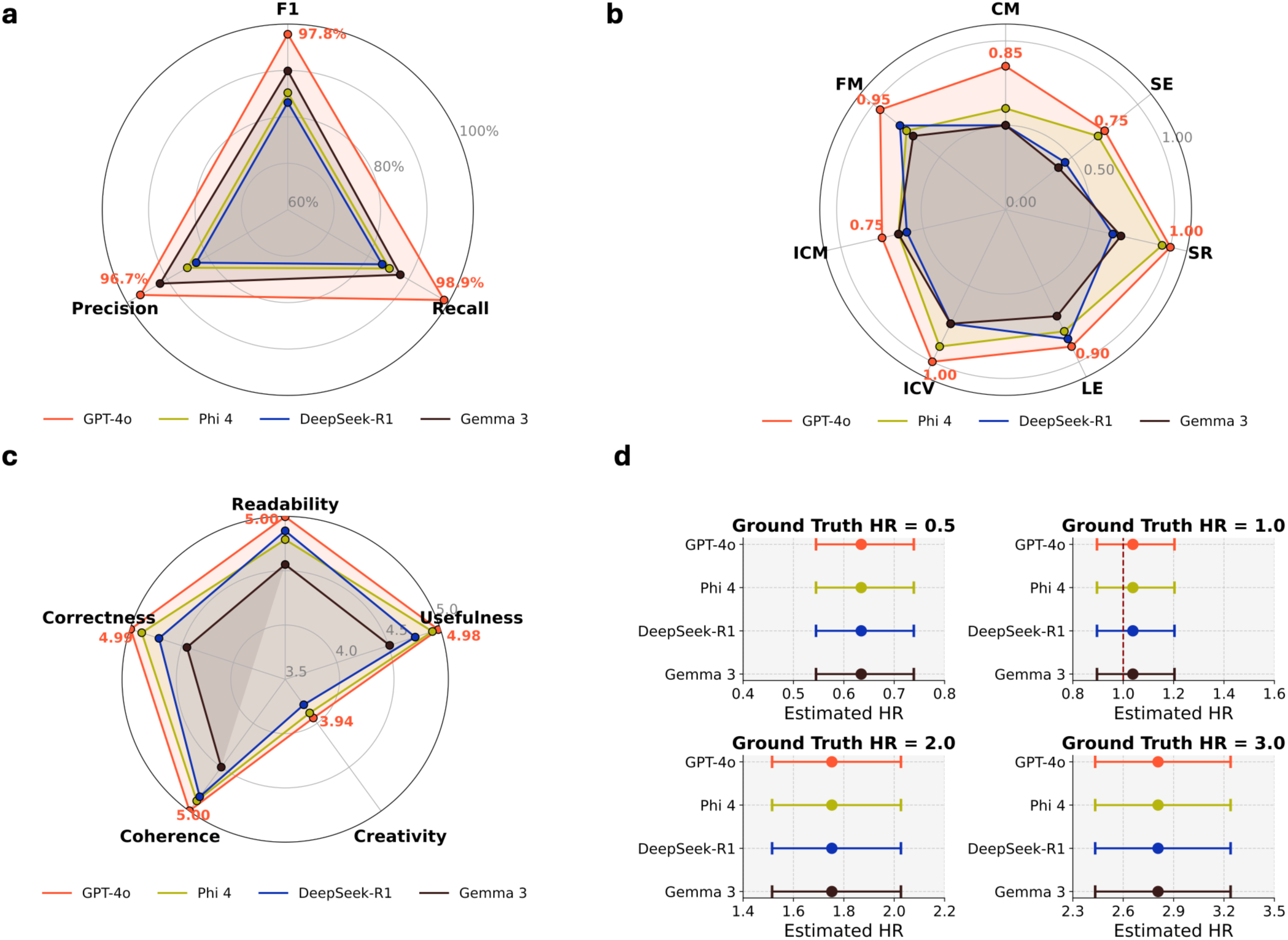
Evaluation of the agents in EmulatRx. (a) Comparison of entity parsing performance among four LLMs: GPT-4o, Phi-4, DeepSeek-R1, and Gemma 3, using precision, recall, and F1-score. GPT-4o achieved the highest recall (98.9%) and precision (96.7%), while Gemma 3 demonstrated the lowest performance. (b) Accuracy of SQL generation by the Informatician across seven categories of common error types, including Concept Missing (CM), Functional Misuse (FM), Incorrect Concept Mapping (ICM), Integrity Constraint Violation (ICV), Logic Error (LE), Schema Reference Error (SR), and Syntax Error (SE). Accuracy is defined as the proportion of trials in which no error of a given type occurred. GPT-4o consistently achieves the highest accuracy across all error categories, with particularly strong performance in logic consistency (LE), schema referencing (SR), and integrity constraint handling (ICV), indicating its ability to preserve both semantic fidelity and structural validity when translating complex eligibility criteria into executable SQL queries. (c) Evaluation of Clinician agent-generated responses based on readability, correctness, coherence, creativity, and usefulness. GPT-4o consistently outperformed other models, achieving the highest scores across all dimensions. (d) Performance comparison of estimated hazard ratios (HR) against ground truth HR values (0.5, 1.0, 2.0, 3.0) of the Statistician agent. All agents produced estimates closest to the ground truth across all scenarios.

### Evaluations on Informatician

For the Informatician, evaluations focus on the SQL generation following Criteria2Query3.0,^30^ using all clinical trials parsed by Trialist. Each generated SQL query is reviewed to identify potential errors manually. To ensure query correctness, the generated SQL statements were executed on the database, and their output was compared with expected cohort retrieval results.

Errors identified in the SQL queries were classified into seven categories (Supplementary Table 2), broadly falling into two types: semantic errors and structural errors.^30^ Semantic errors included logic errors, where relational, temporal, or numerical expressions were misinterpreted (e.g., incorrect handling of age restrictions), concept omissions, where extracted clinical concepts were not incorporated into the query, and incorrect concept mapping, where the linkage between extracted criteria and database standard terminologies was inaccurate. Structural errors encompassed function misuse, integrity constraint violations, schema reference errors, and syntax errors, which affected the overall execution and validity of the queries.

The results are summarized in Figure 4b, where accuracy is defined as the proportion of trials in which no error of a given type occurred. We observed that GPT-4o achieved the highest accuracy across all error categories, indicating superior reliability in translating trial specifications into executable SQL queries. In particular, GPT-4o demonstrated strong performance in preserving logical consistency and schema validity, while maintaining high accuracy across both semantic and structural error types. Phi-4 showed moderate accuracy, whereas DeepSeek-R1 and Gemma 3 consistently exhibited lower accuracy, especially for error types related to concept grounding and query syntax. These results indicate that GPT-4o is the most reliable model for SQL generation within the Informatician agent.

The evaluation results using GPT-4o further revealed a significant association between total error count and eligibility criteria complexity (Table 2). We adopted a heuristic definition from prior literature^73^ to classify eligibility criteria as simple or complex and quantified overall complexity of one trial using a complexity score, defined as the proportion of complex eligibility criteria. We then examined the relationship between total error count and eligibility criteria complexity, observing a statistically significant positive correlation (Spearman’s ρ = 0.45, p-value = 0.043<0.05).^74^ This result indicates that higher eligibility criteria complexity is associated with increased error count. This trend suggests that query accuracy degrades as eligibility criteria become more complex, reflecting the growing difficulty of accurately parsing and translating intricate clinical conditions into executable SQL queries. Overall, the Informatician agent demonstrates strong SQL generation capabilities. These findings are consistent with prior observations reported in Criteria2Query 3.0.^30^

**Table 2.** Complexity of eligibility criteria and total error count across all error types. #Simple ECs denotes the number of simple eligibility criteria, whereas #Complex ECs denotes the number of complex eligibility criteria. A criterion was considered “simple” if it expressed a single clinical concept (or its negation) or a single quantitative comparison, and could be rewritten in such a form without changing its meaning. Criteria involving Boolean logic, temporal constraints, conditional statements, or multiple interacting clinical concepts were classified as “complex”.^73^ Comp. Score indicates the eligibility criteria complexity score. #Errors represents the total number of errors across all error types.

| Trial ID | #EC | #Simple EC | #Complex EC | Comp. Score | #Errors | Trial ID | #EC | #Simple EC | #Complex EC | Comp. Score | #Errors |
| --- | --- | --- | --- | --- | --- | --- | --- | --- | --- | --- | --- |
| Acute diseases on MIMIC-IV data |  |  |  |  |  | Chronic diseases on INSIGHT |  |  |  |  |  |
| NCT00475852 | 7 | 3 | 4 | 0.57 | 3 | NCT04691505 | 6 | 1 | 5 | 0.83 | 3 |
| NCT02856698 | 4 | 3 | 1 | 0.25 | 1 | NCT05457855 | 6 | 2 | 4 | 0.67 | 4 |
| NCT03872011 | 11 | 4 | 7 | 0.64 | 5 | NCT05635357 | 6 | 2 | 4 | 0.67 | 0 |
| NCT04134403 | 10 | 7 | 3 | 0.3 | 1 | NCT05768945 | 7 | 2 | 5 | 0.71 | 3 |
| NCT06091982 | 5 | 3 | 2 | 0.4 | 2 | NCT05794997 | 7 | 2 | 5 | 0.71 | 2 |
| NCT03258684 | 5 | 4 | 1 | 0.2 | 0 | NCT05611372 | 11 | 4 | 7 | 0.64 | 0 |
| NCT03592693 | 10 | 4 | 6 | 0.6 | 2 | NCT05457868 | 6 | 2 | 4 | 0.67 | 1 |
| NCT01560650 | 8 | 4 | 4 | 0.5 | 0 | NCT03659682 | 16 | 6 | 10 | 0.63 | 0 |
| NCT01223690 | 8 | 3 | 5 | 0.625 | 0 | NCT01723228 | 13 | 4 | 9 | 0.69 | 2 |
| NCT00953303 | 10 | 5 | 5 | 0.5 | 1 | NCT01313845 | 11 | 3 | 8 | 0.73 | 3 |

For the emulated trial cohort of NCT03872011 with the complexity score of 0.64,^73^ EmulatRx identified 33 additional eligible patients through natural language processing of clinical notes that were not discoverable via structured electronic health record data alone (Supplementary Figure 1), highlighting the added sensitivity afforded by unstructured data analysis.

### Evaluations on Statistician

For the Statistician, we primarily evaluate the causal inference methods. Covariate balance is measured using standardized mean differences (SMD),^75^ aiming for values below 0.1. The accuracy of survival estimates is compared against published literature, while consistency with known trial results is assessed. Generated reports are also reviewed for clarity and alignment with research objectives. We also performed several evaluations of the various components of the Statistician using synthetic datasets.

For the first evaluation, we created a synthetic dataset with 1,000 subjects and 10 covariates sampled from a standard normal distribution. Treatment assignment was imbalanced using a logistic function based on Sequential Organ Failure Assessment (SOFA) and age. SOFA is a tool used to assess organ dysfunction in critically ill patients in the ICU.^76^ Survival times were simulated under an exponential proportional hazards model with a baseline hazard of 0.1 and treatment effects corresponding to ground truth HRs of 0.5, 1, 2, and 3. Random censoring was incorporated using an exponential distribution, and the ground truth average treatment effect (ATE) was defined as the risk difference at a 10-time unit horizon. Four different large language model (LLM) bases (GPT-4o, Phi-4, DeepSeek-R1, and Gemma 3) were used to drive agent decision-making and model selection (Table 3).

**Table 3.** Evaluation of Statistician’s ability to recapitulate ground truth effect sizes from synthetic datasets.

| LLM | Balancing Method | Chosen Outcome Models | Estimated HR | Estimated CI | Ground Truth HR | Estimated ATE | Ground Truth ATE |
| --- | --- | --- | --- | --- | --- | --- | --- |
| GPT-4o | Propensity Score Matching (PSM) | Cox Proportional Hazards, Kaplan-Meier Estimator | 0.6345 | (0.5448, 0.7389) | 0.5 | -0.0997 | -0.175934 |
| Phi-4 | Propensity Score Matching (PSM) | Cox Proportional Hazards, Random Survival Forest | 0.6345 | (0.5448, 0.7389) | 0.5 | -0.0997 | -0.175934 |
| DeepSeek-R1 | Propensity Score Matching (PSM) | Cox Proportional Hazards, Random Survival Forests | 0.6345 | (0.5448, 0.7389) | 0.5 | -0.0997 | -0.175934 |
| Gemma 3 | Propensity Score Matching (PSM) | Cox Proportional Hazards Regression | 0.6345 | (0.5448, 0.7389) | 0.5 | -0.0997 | -0.175934 |
| GPT-4o | Propensity Score Matching (PSM) | Cox Proportional Hazards, Kaplan-Meier Estimator | 1.0383 | (0.8958, 1.2034) | 1 | 0.0244 | 0 |
| Phi-4 | Propensity Score Matching (PSM) | Cox Proportional Hazards, Random Survival Forest | 1.0383 | (0.8958, 1.2034) | 1 | 0.0244 | 0 |
| DeepSeek-R1 | Propensity Score Matching (PSM) | Cox Proportional Hazards, Random Survival Forests | 1.0383 | (0.8958, 1.2034) | 1 | 0.0244 | 0 |
| Gemma 3 | Propensity Score Matching (PSM) | Cox Proportional Hazards Regression | 1.0383 | (0.8958, 1.2034) | 1 | 0.0244 | 0 |
| GPT-4o | Propensity Score Matching (PSM) | Cox Proportional Hazards, Kaplan-Meier Estimator | 1.7524 | (1.5152, 2.0267) | 2 | 0.1461 | 0.16326836 |
| Phi-4 | Propensity Score Matching (PSM) | Cox Proportional Hazards, Random Survival Forest | 1.7524 | (1.5152, 2.0267) | 2 | 0.1461 | 0.16326836 |
| DeepSeek-R1 | Propensity Score Matching (PSM) | Cox Proportional Hazards, Random Survival Forests | 1.7524 | (1.5152, 2.0267) | 2 | 0.1461 | 0.16326836 |
| Gemma 3 | Propensity Score Matching (PSM) | Cox Proportional Hazards Regression | 1.7524 | (1.5152, 2.0267) | 2 | 0.1461 | 0.16326836 |
| GPT-4o | Propensity Score Matching (PSM) | Cox Proportional Hazards, Kaplan-Meier Estimator | 2.8074 | (2.4330, 3.2394) | 3 | 0.2225 | 0.28317249 |
| Phi-4 | Propensity Score Matching (PSM) | Cox Proportional Hazards, Random Survival Forest | 2.8074 | (2.4330, 3.2394) | 3 | 0.2225 | 0.28317249 |
| DeepSeek-R1 | Propensity Score Matching (PSM) | Cox Proportional Hazards, Random Survival Forests | 2.8074 | (2.4330, 3.2394) | 3 | 0.2225 | 0.28317249 |
| Gemma 3 | Propensity Score Matching (PSM) | Cox Proportional Hazards Regression | 2.8074 | (2.4330, 3.2394) | 3 | 0.2225 | 0.28317249 |

Across all LLMs, propensity score matching was consistently chosen as the balancing method by the Statistician. For a ground truth HR of 0.5, the Statistician estimated a HR of 0.6345 (95% CI: 0.5448–0.7389) and an ATE of –0.0997, compared with a ground truth ATE of –0.1759. When the true effect was null (HR = 1), the estimated HR was 1.0383 (95% CI: 0.8958–1.2034) with an ATE of 0.0244 (ground truth ATE = 0). For a moderate effect (HR = 2), the estimated HR was 1.7524 (95% CI: 1.5152–2.0267) and the ATE was 0.1461 (ground truth ATE = 0.1633). Finally, for HR = 3, the Statistician produced an estimated HR of 2.8074 (95% CI: 2.4330–3.2394) with an ATE of 0.2225, versus a ground truth ATE of 0.2832. These estimates were robust across all LLMs and outcome model selections, demonstrating that the Statistician can reliably recapitulate the ground truth parameters in trial emulation. It is important to note that the HRs and confidence intervals in Table 3 are identical across different LLMs. This is because of the Statistician agent’s tool-use architecture. The agent’s role is to select the optimal statistical method based on data characteristics, while the actual computation is done by deterministic statistical libraries. In this evaluation, all four LLMs identified the same optimal strategy for the synthetic dataset. As such, they used the exact same execution tools, leading to identical numerical results.

In the second evaluation, we generated a synthetic survival dataset with 1,000 subjects containing two covariates (SOFA and age) and a binary treatment. In the overall (unstratified) analysis, the treatment effect was engineered such that the average HR was null (i.e., no significant treatment effect). However, the treatment effect was designed to interact with the SOFA score such that, when subjects were grouped by a clinically meaningful cutoff, significant subgroup effects would emerge. Specifically, for treated subjects, the log hazard effect was set to +1 when the SOFA score was below a threshold (e.g., <8.0) and –1 when the score was above that threshold; a constant was subtracted to force the marginal (unstratified) HR to be 1. We evaluated the ability of our multi-agent framework (incorporating Statistician and Clinician agents) to (1) detect the absence of an overall treatment effect and (2) uncover significant subgroup effects by stratifying on SOFA. Three different large language model (LLM) bases (GPT-4o, Phi-4, and DeepSeek-R1) were deployed to drive agent decisions regarding subgroup creation and subsequent survival analysis using Cox proportional hazards models.

Table 4 shows the results. Unadjusted Cox modeling of the full dataset revealed no significant treatment effect. However, after stratification by SOFA, subgroup analyses produced statistically significant differences. For analyses driven by GPT-4o and Phi-4, subjects with “SOFA < 8.0” had an estimated HR of 1.3849 [95% CI: 1.2101, 1.5850] while those with “SOFA ≥ 8.0” had an estimated HR of 0.7280 [95% CI: 0.5921, 0.8950]. In contrast, the DeepSeek-R1 and Gemma 3 driven analysis identified a slightly different cutoff (SOFA < 7.0 vs. SOFA ≥ 7.0) yielding HRs of 1.3443 [95% CI: 1.1760, 1.5366] and 0.8286 [95% CI: 0.6853, 1.0019], respectively. These findings demonstrate that while the overall analysis recapitulated the engineered null effect, appropriate stratification by the effect modifier SOFA uncovered significant heterogeneity in treatment response. The consistency across different LLM bases supports the robustness of our multi-agent evaluation framework.

**Table 4.** Evaluation of Statistician’s ability to recapitulate ground truth effect sizes after subgroup stratification.

| LLM | Number of Subgroup Analyses Done | Significant Stratification Levels | Estimated HR | Ground Truth HR | Estimated CI |
| --- | --- | --- | --- | --- | --- |
| GPT-4o | 1 | SOFA < 8.0<br>SOFA ≥ 8.0 | 1.3849<br>0.7280 | 1.4521<br>0.6473 | (1.2101, 1.5850)<br>(0.5921, 0.8950) |
| Phi-4 | 2 | SOFA < 8.0<br>SOFA ≥ 8.0 | 1.3849<br>0.7280 | 1.4521<br>0.6473 | (1.2101, 1.5850)<br>(0.5921, 0.8950) |
| DeepSeek-R1 | 1 | SOFA < 7.0<br>SOFA ≥ 7.0 | 1.3443<br>0.8286 | 1.4060<br>0.7564 | (1.1760, 1.5366)<br>(0.6853, 1.0019) |
| Gemma 3 | 1 | SOFA < 7.0<br>SOFA ≥ 7.0 | 1.3443<br>0.8286 | 1.4060<br>0.7564 | (1.1760, 1.5366)<br>(0.6853, 1.0019) |

We evaluated our EC optimization algorithm by creating synthetic datasets with known ground-truth importances for each EC and then measuring how closely the Monte Carlo-based Shapley estimator recovered these importances under varying numbers of ECs. For each run, we drew a specified number of ECs (from 2 to 20), sampled true importances uniformly from a predefined range, and combined these with a baseline hazard-ratio (HR) value. We enumerated every possible subset of the ECs and generated noisy HRs by adding Gaussian noise (standard deviation = 0.5) to each subset’s baseline plus its summed importances. Our algorithm then performed randomized permutations of the ECs to compute incremental differences in HR from the empty to the full set of rules, up to a maximum of 1000 iterations or until the standard error of the mean (SEM) reached a small tolerance (1e-3). We compared each criterion’s estimated Shapley value against its ground-truth importance using mean absolute error (MAE). Supplementary Table 3 shows the MAE for varying number of ECs.

Supplementary Table 3 shows that, even in the presence of moderate noise, the algorithm converges effectively and typically produces low MAE values, indicating good agreement between the estimated Shapley values and the known true importances.

To demonstrate the utility of adaptive sample size calculation, we present a showcase based on trial NCT00475852, which investigated the effects of nesiritide in patients with acute heart failure. In the real-world cohort assembled by the Informatician agent from the MIMIC-IV database, a total of 6,971 patients met the eligibility criteria aligned with the original trial protocol. However, to assess the feasibility of prospective trial planning and explore whether fewer participants would suffice to achieve adequate statistical power, the Statistician agent was prompted to initiate sample size estimation. Using the Schoenfeld formula for the Cox proportional hazards model, the agent incorporated key characteristics from the observed data, including treatment allocation ratio, baseline event rate, censoring proportion, and average follow-up time. The target specifications for the calculation were a Type I error rate (α) of 0.05, power (1–β) of 0.80, and an expected HR based on prior trial literature. The automated estimation yielded a required sample size of 3,107 patients, largely lower than the original cohort size of 6,971. This reduction reflects the relatively high event rate and long follow-up observed in the RWD, which collectively increased the statistical efficiency of the emulated trial design. This result highlights the value of data-driven sample size planning within EmulatRx. By dynamically adjusting for empirical characteristics of the study population, the system can help optimize resource allocation while preserving inferential power. Such functionality is especially valuable in real-world CTD scenarios with limited resources.

### Evaluations on Clinician

The Clinician agent is evaluated for its ability to integrate medical expertise into the workflow. A questionnaire based on a 5-point Likert scale^77^ was provided to three independent clinical domain experts to assess the relevance, clarity, and accuracy of the Clinician’s recommendations.^30^ Additionally, the agent’s efficiency in refining trial designs and resolving discrepancies is measured through qualitative feedback from domain experts.

- Figure 4c presents the evaluation of Clinician-generated responses across five key dimensions: readability, correctness, coherence, creativity, and usefulness, with detailed subcategory scores provided in Table 5. Overall, GPT-4o outperformed all other models, achieving the highest average score of 4.88. Phi-4 and DeepSeek-R1 followed with strong overall performance (4.78 and 4.71, respectively), while Gemma 3 lagged with a significantly lower average score of 4.40.<u>Readability</u>: GPT-4o achieved near-perfect consistency in readability, scoring 4.99–5.00 across all aspects, including writing clarity, logical structure, and appropriate terminology. DeepSeek-R1 followed with solid scores around 4.86–4.88, showing well-organized responses. Phi-4 scored slightly lower (4.75–4.82), suggesting occasional inconsistencies in formatting or flow. In contrast, Gemma 3 underperformed in this dimension, with scores ranging from 4.54 to 4.58, indicating relative challenges in producing clearly formatted and accessible clinical content.
- <u>Correctness</u>: -GPT-4o again led in this category, scoring 4.99 for factual accuracy and 4.99–5.00 in logical reasoning and domain-specific usage, reflecting both precision and reasoning strength. Phi-4 was close behind (4.88–4.89), demonstrating high reliability. DeepSeek-R1 showed slightly lower performance (4.71–4.73), pointing to occasional gaps in factual or logical precision^79^.. Gemma 3 trailed with scores between 4.42 and 4.48, underscoring its relative limitation in maintaining strict clinical accuracy.
- <u>Coherence</u>: GPT-4o performed strongly again (4.99–5.00), maintaining logical consistency across individual and multiple responses. Phi-4 showed robust coherence (4.86–4.93), effectively avoiding contradictions. DeepSeek-R1 also produced coherent outputs (4.83–4.84), while Gemma 3’s scores (4.47–4.53) pointed to comparatively greater structural fragility and potential internal inconsistencies. <u>Creativity</u>: All four models showed room for growth in this aspect. GPT-4o displayed relatively higher creativity (3.93-3.95), especially in offering multiple perspectives. <u>Usefulness</u>: GPT-4o stood out as the most practically useful model, scoring 4.97–4.99 for application fit and modifiability. Phi-4 was highly competitive in this category (4.90–4.94), indicating strong utility for downstream tasks. DeepSeek-R1 scored slightly lower (4.74–4.77), while Gemma 3 scored the lowest (4.50–4.52), indicating its output may require more refinement for practical application.

**Table 5:** Questionnaire and mean scores on Clinician-generated responses across three participants. P-values were determined using Kruskal Wallis test^78^.

| Category | Criterion | GPT-4o | Phi-4 | Deepseek-R1 | Gemma 3 | P-Value | Best Model |
| --- | --- | --- | --- | --- | --- | --- | --- |
| Readability | The overall writing format is easy to understand. | 5.00 | 4.82 | 4.86 | 4.58 | < 0.001 | GPT-4o |
|  | The response is well-structured and logically organized (e.g., proper use of paragraphs, bullet points). | 5.00 | 4.75 | 4.86 | 4.54 | < 0.001 | GPT-4o |
|  | The terminology and language style are appropriate for the intended audience. | 4.99 | 4.79 | 4.88 | 4.55 | < 0.001 | GPT-4o |
| Correctness | The response accurately conveys factual information and is free from hallucinations. | 4.99 | 4.89 | 4.71 | 4.42 | < 0.001 | GPT-4o |
|  | The logical reasoning in the response is sound and follows expected principles. | 4.99 | 4.88 | 4.73 | 4.45 | < 0.001 | GPT-4o |
|  | The response correctly uses technical or domain-specific terms when applicable. | 5.00 | 4.89 | 4.72 | 4.48 | < 0.001 | GPT-4o |
| Coherence | The response maintains internal logical consistency throughout. | 4.99 | 4.86 | 4.84 | 4.50 | < 0.001 | GPT-4o |
|  | If multiple responses are generated for the same prompt, they are consistent with each other. | 5.00 | 4.86 | 4.83 | 4.47 | < 0.001 | GPT-4o |
|  | The response avoids contradictions or conflicting statements. | 5.00 | 4.93 | 4.84 | 4.53 | < 0.001 | GPT-4o |
| Creativity | The response demonstrates originality and innovative reasoning. | 3.93 | 3.90 | 3.80 | 3.47 | < 0.001 | GPT-4o |
|  | The answer provides novel insights beyond common knowledge. | 3.95 | 3.87 | 3.78 | 3.53 | < 0.001 | GPT-4o |
|  | The response presents multiple perspectives when appropriate. | 3.94 | 3.88 | 3.79 | 3.47 | < 0.001 | GPT-4o |
| Usefulness | The response is useful for the intended application (e.g., SQL generation, summarization, question answering). | 4.97 | 4.93 | 4.76 | 4.51 | < 0.001 | GPT-4o |
|  | The generated response is easy to modify or refine for further use. | 4.99 | 4.90 | 4.74 | 4.52 | < 0.001 | GPT-4o |
|  | The response format aligns with the requirements of the target application. | 4.98 | 4.94 | 4.77 | 4.50 | < 0.001 | GPT-4o |
| Average Score |  | 4.88 | 4.78 | 4.71 | 4.40 | < 0.001 | GPT-4o |

Finally, we conducted an ablation study to validate the contribution of the RAG module. The results demonstrated that RAG is essential for knowledge grounding; while the non-RAG baseline correctly reported statistical outputs, the RAG-enabled agent produced significantly richer summaries that contextualized findings against existing biomedical literature and consistently included granular metrics such as Average Treatment Effects. In summary, GPT-4o delivered the most accurate, relevant, and clinically useful responses, making it the clear leader among all evaluated LLMs. DeepSeek-R1 and Phi-4 demonstrated decent usability with minor flaws, while Gemma 3 consistently underperformed across nearly all dimensions, indicating that its clinical reasoning and formatting still require significant improvement for integration in expert workflows.

### Showcases

To demonstrate how EmulatRx operates on diverse clinical questions, we present three separate disease-based examples using GPT-4o, each reflecting distinct trial emulation scenarios (Figure 5). These examples illustrate how the Supervisor, Trialist, Informatician, Clinician, and Statistician agents coordinate to parse protocols, handle eligibility criteria, select covariates, balance confounders, and produce final analyses.

**Figure 5.**
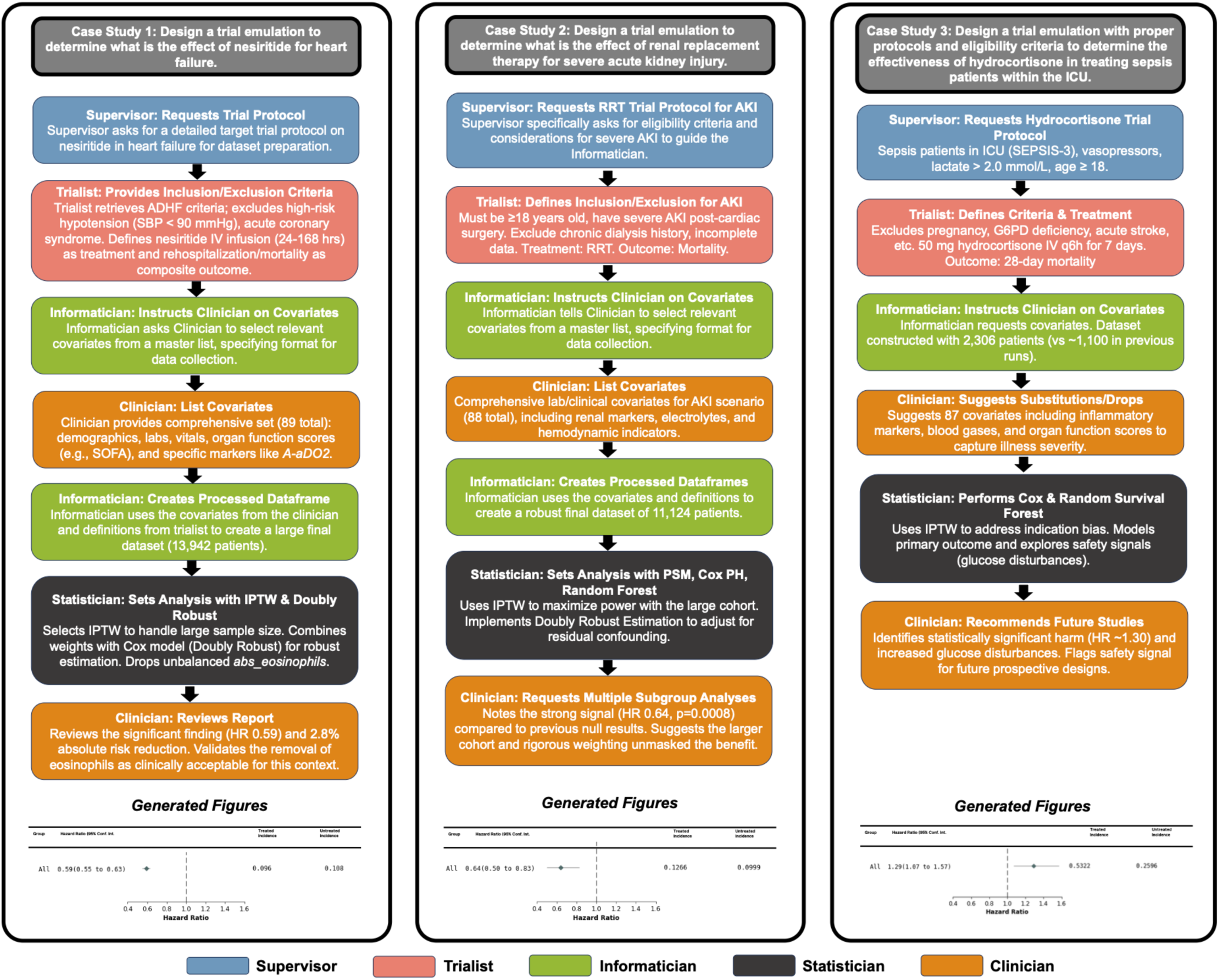
Showcases on three clinical trials. Text within the worflows are generated by using GPT-4o to summarize EmulatRx logs for each case study.

### Case 1: Impact of Nesiritide in Acute Heart Failure Patients

In this first demonstration, EmulatRx emulated a clinical trial (NCT00475852) assessing nesiritide’s impact on heart failure outcomes. The Supervisor initiated the workflow by requesting a detailed target trial protocol. The Trialist agent retrieved and parsed the inclusion criteria, specifying patients hospitalized for acute decompensated heart failure (ADHF) or diagnosed within 48 hours, while strictly excluding those with high-risk hypotension (systolic BP < 90 mmHg), acute coronary syndromes, or specific structural heart defects.

The workflow then moved to a collaborative design phase between the Clinician and Informatician agents. The Clinician agent, leveraging its biomedical knowledge base, proposed a high-dimensional feature space comprising 89 distinct covariates, including granular metrics such as A-aDO2, methemoglobin, and abs_eosinophils alongside standard vital signs and organ function scores (e.g., SOFA). The Informatician agent translated these definitions into OMOP-compliant SQL queries, successfully instantiating a large-scale cohort of 13,942 patients.

During the statistical analysis design, the Statistician agent evaluated the dataset’s properties and selected IPTW over PSM to maximize data utilization given the large sample size. In the covariate balancing phase, the agent detected persistent imbalances in abs_eosinophils. Through an autonomous feedback loop, the Statistician consulted the Clinician, who advised dropping the eosinophil count as it was not a primary confounder for hemodynamic outcomes in this context, allowing the model to achieve satisfactory balance (SMD < 0.1).

The outcome analysis was conducted using a Doubly Robust Estimation framework, combining the IPTW weights with a Cox Proportional Hazards model. The analysis yielded a HR of 0.5913 (95% CI: 0.55–0.63), indicating a statistically significant reduction in the composite risk of rehospitalization and all-cause mortality for the nesiritide-treated group. This result, which aligns with the directional findings of the original ASCEND-HF trial but with a stronger effect size likely due to the specific real-world subpopulation captured, was synthesized into a final report by the Supervisor. The report highlighted a 2.8% absolute risk reduction, concluding that nesiritide offers a quantifiable therapeutic benefit in this observational cohort.

### Case 2: Effect of Renal Replacement Therapy for Severe Acute Kidney Injury

In the second case study, EmulatRx was tasked with emulating a study (NCT06091982) to determine the impact of Renal Replacement Therapy (RRT) on mortality in patients with severe Acute Kidney Injury (AKI). The Supervisor directed the Trialist agent to parse the protocol, which specified inclusion criteria for adults (age >= 18 years) with confirmed severe AKI, while excluding those with prior chronic dialysis history or incomplete baseline data.

The Informatician agent, collaborating with the Clinician to map clinical concepts to the MIMIC-IV schema, successfully identified a robust cohort of 11,124 patients. The Clinician agent ensured the feature space was comprehensive, selecting 88 covariates that included renal function markers, electrolyte panels, and hemodynamic indicators.

Given the substantial sample size, the Statistician agent autonomously selected IPTW as the optimal balancing strategy to preserve the full statistical power of the cohort, rather than pruning samples via matching. The agent implemented a Doubly Robust Estimation method, adjusting for residual confounding in the outcome model. In contrast to earlier iterations that reported null findings, this high-fidelity emulation revealed a HR of 0.6443 (p=0.0008), suggesting a significant survival benefit associated with RRT in this critically ill population. The final report synthesized these findings, noting that the rigorous adjustment for confounding provided by the IPTW approach likely unmasked a treatment effect that was previously obscured in smaller, less representative subsamples.

### Case 3: Effect of Hydrocortisone in Septic Shock Patients

In the final example, EmulatRx emulated a trial (NCT04134403) evaluating hydrocortisone therapy in patients with septic shock. The Supervisor requested a protocol targeting patients with persistent vasopressor dependence and lactate levels >2.0 mmol/L. The Trialist agent parsed the eligibility criteria to exclude patients with confounding conditions such as major hemorrhage, burns, or prolonged vasopressor use prior to randomization.

Working in tandem, the Clinician and Informatician agents curated a dataset of 2,306 patients, a substantial increase over the 1,153 patients identified in the previous run. The Clinician agent specified 87 covariates, including granular inflammatory markers and organ function scores, ensuring a dense feature representation for causal inference.

The Statistician agent employed IPTW coupled with Doubly Robust Estimation. This emulation detected a Hazard Ratio of 1.2983 (95% CI: 1.07–1.57; p=0.0082), indicating a statistically significant increased risk of 28-day mortality in the hydrocortisone-treated group within this specific MIMIC-IV subpopulation. These findings are consistent with previous analyses on the MIMIC-IV data, as it relates to the usage of steroids on sepsis patients^8^. The final report flagged this finding as a critical safety signal, prompting the Clinician agent to recommend extreme caution and further subgroup stratification in future prospective designs. This case underscores EmulatRx’s capacity to identify potential harms in real-world settings that may differ from controlled trial environments due to population heterogeneity.

We further examined the interaction dynamics to assess whether our framework indeed operates as a collaborative multi-agent framework. Supplementary Figure 2 summarizes the observed agent-to-agent interaction transitions aggregated across all trials and LLMs. The patterns illustrate rich inter-agent interactions, including conversations between the Informatician and Clinician during cohort construction, between the Statistician and Clinician during covariate and model refinement, and coordination mediated by the Supervisor. These observations indicate that our framework does not function as a simple linear pipeline, but rather as a dynamically interacting system in which agents exchange intermediate results and feedback to iteratively refine trial design and analysis.

To quantitatively evaluate the efficiency of the framework, we measured the end-to-end execution time of the full trial design pipeline. Supplementary Figure 3 reports the wall-clock runtime across executions with all trials among different LLM backends. We observed that GPT-4o achieves the shortest median runtime (5.75±1.52 minutes). Phi-4 and Gemma-3 exhibit moderate runtimes (20.95±7.94 minutes, 25.87±4.38 minutes, respectively), while DeepSeek-R1 shows higher variance and longer runtimes (31.36±14.51 minutes), largely attributable to its extended chain-of-thought reasoning and local inference latency. In contrast, manual execution of comparable workflows requiring domain experts to translate trial protocols, write and debug SQL queries, engineer covariates, conduct causal analyses, and iteratively refine design choices typically spans multiple days to weeks.^80–83^ These results provide empirical evidence that EmulatRx not only facilitates but substantially accelerates the clinical trial design process.

## Discussions

EmulatRx represents an advancement in the use of agentic systems for empowering CTD. By integrating role-specialized LLM agents, EmulatRx transforms a traditionally manual, expertise-intensive and time-consuming CTD process into an autonomous pipeline. Recent studies have explored the use of large language model (LLM)–based agents for clinical trial–related tasks, such as trial outcome prediction, protocol reasoning, and structured clinical decision support. For example, AUTOCT^84^ and ClinicalAgent^85^ employ multi-agent reasoning to support confined subtasks within the clinical trial pipeline and are evaluated using task-specific benchmarks. Related reviews^86^ have further summarized emerging agentic workflows in clinical and translational research. In contrast, EmulatRx targets end-to-end clinical trial design, encompassing protocol construction, cohort instantiation from real-world data, causal effect estimation, and result interpretation.

Among the five agents, the Trialist demonstrated its capability in retrieving and standardizing trial information, achieving a high F1-score of 95.4% using GPT-4o. The Informatician agent translated these parsed elements into SQL queries, showing that LLMs can map trial information into structured cohort definitions. The Statistician agent effectively executed causal inference workflows by matching gold-standard HR estimates across synthetic and RWD. The Clinician agent ensured medical validity and interpretability through structured reasoning grounded in biomedical literature.

Notably, GPT-4o consistently outperformed locally deployed models (Phi-4, DeepSeek-R1, and Gemma 3) across tasks, particularly in logical accuracy, syntactic correctness, and clinical relevance. Our analysis of the Informatician indicates that the performance disparity between GPT-4o and local models (Phi-4, DeepSeek-R1, Gemma 3) stems primarily from differences in reasoning capabilities and instruction adherence rather than context window limitations. The local models struggled significantly with concept mapping and SQL syntax, particularly when facing high-complexity eligibility criteria. This suggests that while off-the-shelf local models currently lack the domain grounding required for reliable OMOP-SQL translation, they could likely be improved via targeted fine-tuning on clinical Text-to-SQL datasets.

By modularizing CTD into distinct roles, i.e., Supervisor, Trialist, Informatician, Clinician, and Statistician, EmulatRx reflects the natural division of efforts in clinical research teams. The architecture enables agents to iterate collaboratively with refining eligibility definitions, proposing surrogate covariates, or adjusting statistical models, which helps mirror the dynamic, interdisciplinary nature of CTD in practice. The integration of Reinforcement Learning from Human Feedback (RLHF) further aligns model behavior with human experts, allowing agents to improve the response quality. These designs allow EmulatRx to function not merely as a pipeline, but as a learning ecosystem that continuously refines both knowledge and execution.

Effect sizes derived by EmulatRx are inherently specific to the underlying real-world dataset and the precise cohort definitions generated during emulation. In many RWE scenarios, a primary analysis may yield a non-significant result. To address this, EmulatRx is designed to look beyond the aggregate treatment effect; the system automatically triggers subgroup analyses and adverse event modeling to uncover heterogeneous effects or safety signals that might otherwise be missed.

From a broader perspective, EmulatRx offers a new paradigm for integrating RWE into clinical research. Traditional pipelines often require substantial human intervention to iteratively reconcile protocol design, making scalability difficult. EmulatRx’s agentic intelligence bridges this gap by autonomously surfacing data issues (e.g., missingness), querying domain knowledge via RAG, and adjusting the trial design accordingly. Such capability is particularly powerful in the domains where real-time insights are essential, and trial feasibility is often constrained.

Several important limitations and directions for future exploration remain. First, there is currently no well-established standardized benchmark for evaluating end-to-end agentic systems for clinical trial design, and evaluations therefore rely on trial-specific analyses, such as alignment with published randomized controlled trials, experiments on synthetic data with known ground truth, and expert clinical assessment. Developing large-scale and unified benchmarks for this problem represents a challenging but important direction for future work.^86–88,87–89^ Second, many clinical trials incorporate not just structured EHR data, but also clinical notes, imaging, genomic profiles, or wearable data. Extending EmulatRx to handle multi-modal inputs would unlock new applications in precision medicine and rare disease research. This would require enhancing the reasoning capabilities of agents like the Clinician and Statistician to synthesize evidence across modalities. Third, although EmulatRx was evaluated on the MIMIC-IV and INSIGHT datasets, its broader applicability across other healthcare systems remains an open question. Extending EmulatRx to function reliably across diverse databases or in federated environments with privacy constraints would enhance its value for multicenter trial design. Forth, the Clinician agent currently relies on RAG over curated literature corpora. Future iterations could enrich this by integrating more biomedical knowledge graphs (e.g., iBKH^90^) to support relational reasoning over structured knowledge. This would enhance the agent’s ability to infer indirect associations (e.g., comorbidities, mechanistic pathways), improve covariate selection, and provide more context-aware recommendations.

## Data

**MIMIC-IV.**^26^ We utilized the Medical Information Mart for Intensive Care (MIMIC-IV) database, a comprehensive critical care repository comprising over 299,000 patients admitted to the Beth Israel Deaconess Medical Center in Boston, MA, between 2008 and 2019. Unlike standard longitudinal repositories, MIMIC-IV provides high-resolution, time-stamped clinical data essential for modeling rapid disease progression in acute care settings. In this work, MIMIC-IV was leveraged to evaluate EmulatRx’s capability to design and emulate trials for acute critical conditions, such as septic shock, acute heart failure, and acute kidney injury, where granular physiological data and free-text clinical narratives are necessary for precise patient matching and outcome assessment.

**INSIGHT.**^27^ We also leveraged the INSIGHT Clinical Research Network (CRN), a large-scale longitudinal EHR repository comprising 5,532,428 patients from five hospital systems across the greater New York City area, with data spanning January 2007 to December 2023. INSIGHT captures longitudinal patient histories across repeated clinical encounters over extended time horizons, enabling the study of chronic disease trajectories. In this work, INSIGHT was used to support clinical trial design and evaluation for chronic conditions, including Alzheimer’s disease and Parkinson’s disease.

## Data Availability

The INSIGHT data can be requested through https://insightcrn.org/. The de-identified data utilized in this study MIMIC-IV can be accessed upon the approval of a formal proposal and the execution of a Data Access Agreement via Physio Net (https://physionet.org/).

## Code Availability

The primary repository is hosted on https://github.com/TrialLab/EmulatRx. All implementation and analysis details are documented within the repository.

## Acknowledgements

The study was not supported by any external funding.

## Author Contributions

F.W. conceived the initial idea. H.L., W.P. and S.R. conceived the method, wrote the codes and performed the computational analysis. H.L., W.P. and S.R. drafted the initial manuscript, with critical revisions by F.W. C.Z. reviewed the manuscript and provided suggestions. F.W. supervised the project. All authors reviewed, provided feedback, and approved the final manuscript.

## Supplemental Materials

**Supplementary Figure 1.**
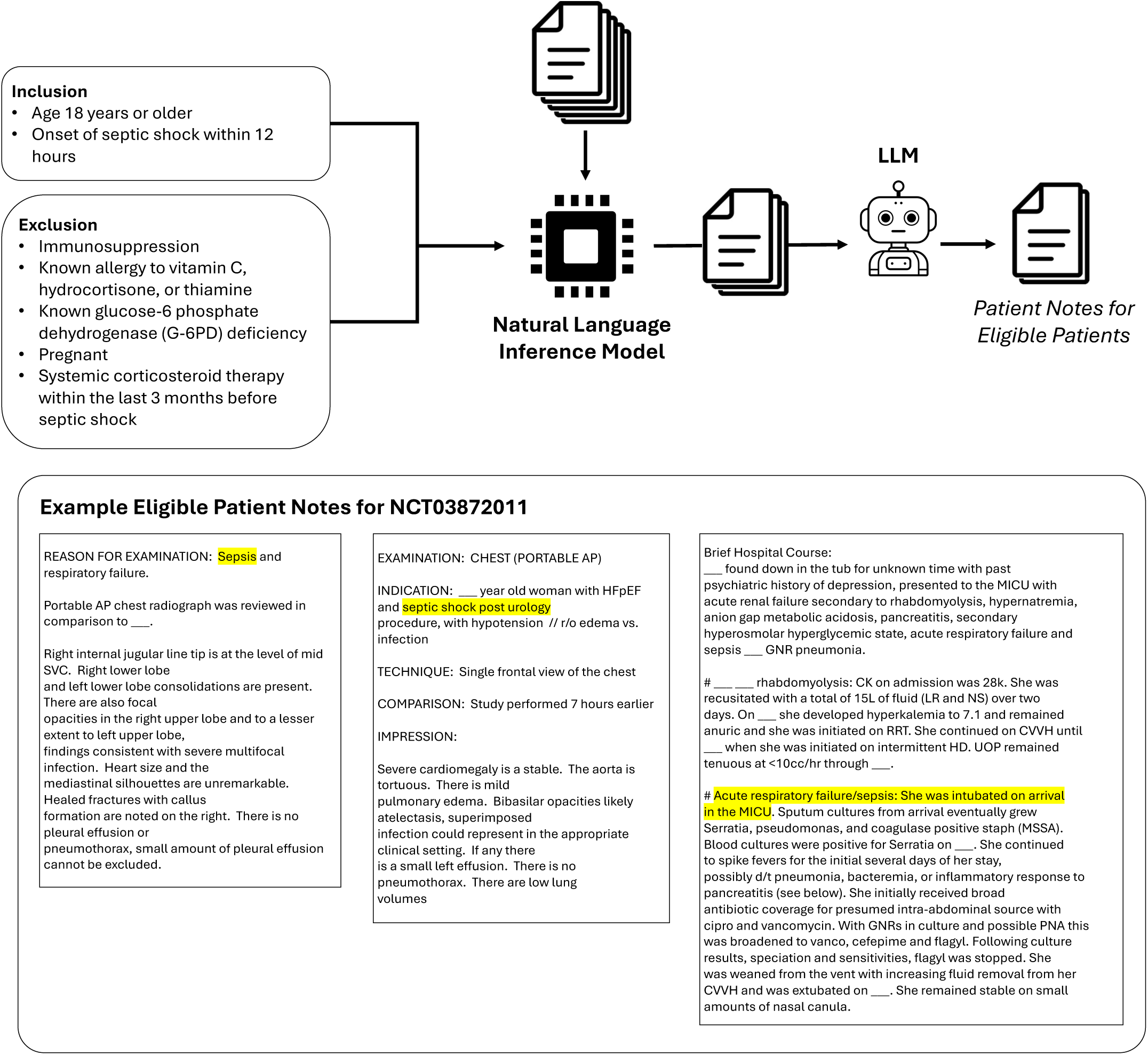
Pipeline of patient matching with unstructured clinical notes. The Informatician agent incorporates NLP techniques to analyze unstructured clinical notes, enriching the patient eligibility assessment process.

**Supplementary Figure 2.**
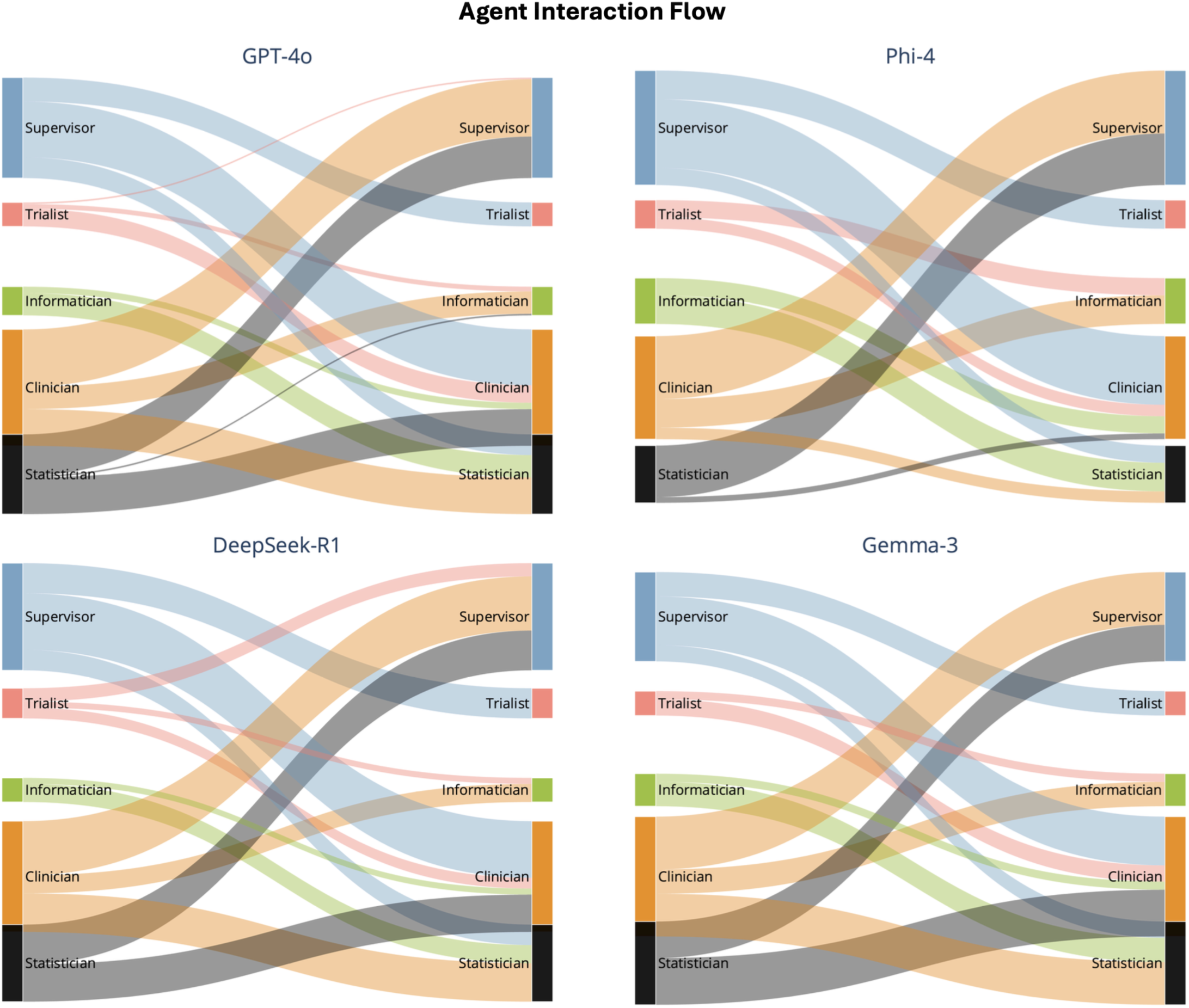
Agent Interaction Flow. Each Sankey diagram visualizes the distribution of interactions between agents when instantiated with different LLMs (GPT-4o, Phi-4, DeepSeek-R1, and Gemma-3). The left and right nodes represent the source and target agents of each interaction step, respectively, and the width of each flow is proportional to the frequency of transitions observed in all trials, illustrating rich and diverse interactions among agents within the framework.

**Supplementary Figure 3.**
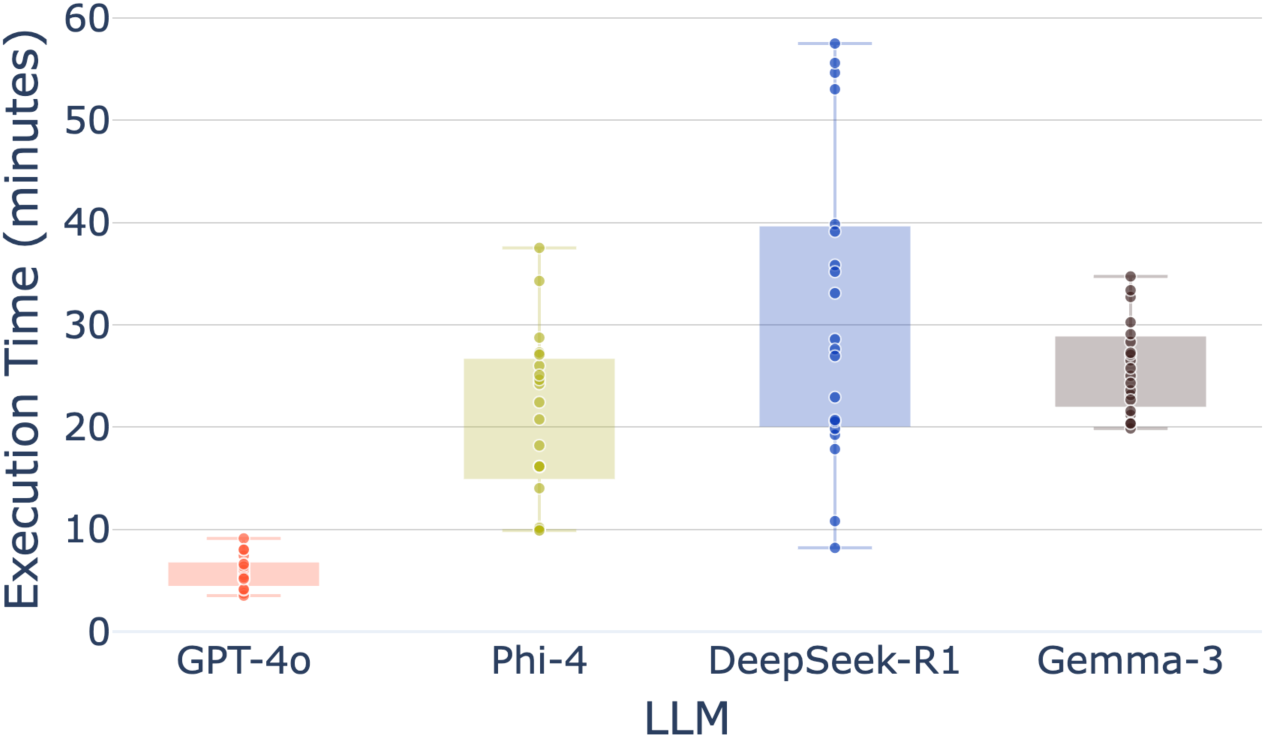
End-to-end execution time of the EmulatRx across different LLM backends. The figure reports runtime (in minutes) for complete trial design runs, obtained on an Apple M1 Pro processor. Each point represents one full execution, and boxplots summarize the distribution across repeated runs. GPT-4o demonstrates the shortest and most stable runtime, while Phi-4, DeepSeek-R1, and Gemma-3 exhibit longer execution times due to local inference overhead. Overall, all configurations complete the end-to-end clinical trial design workflow within minutes, demonstrating substantial acceleration compared to manual expert-driven processes that typically require days to weeks.

**Supplementary Table 1.**
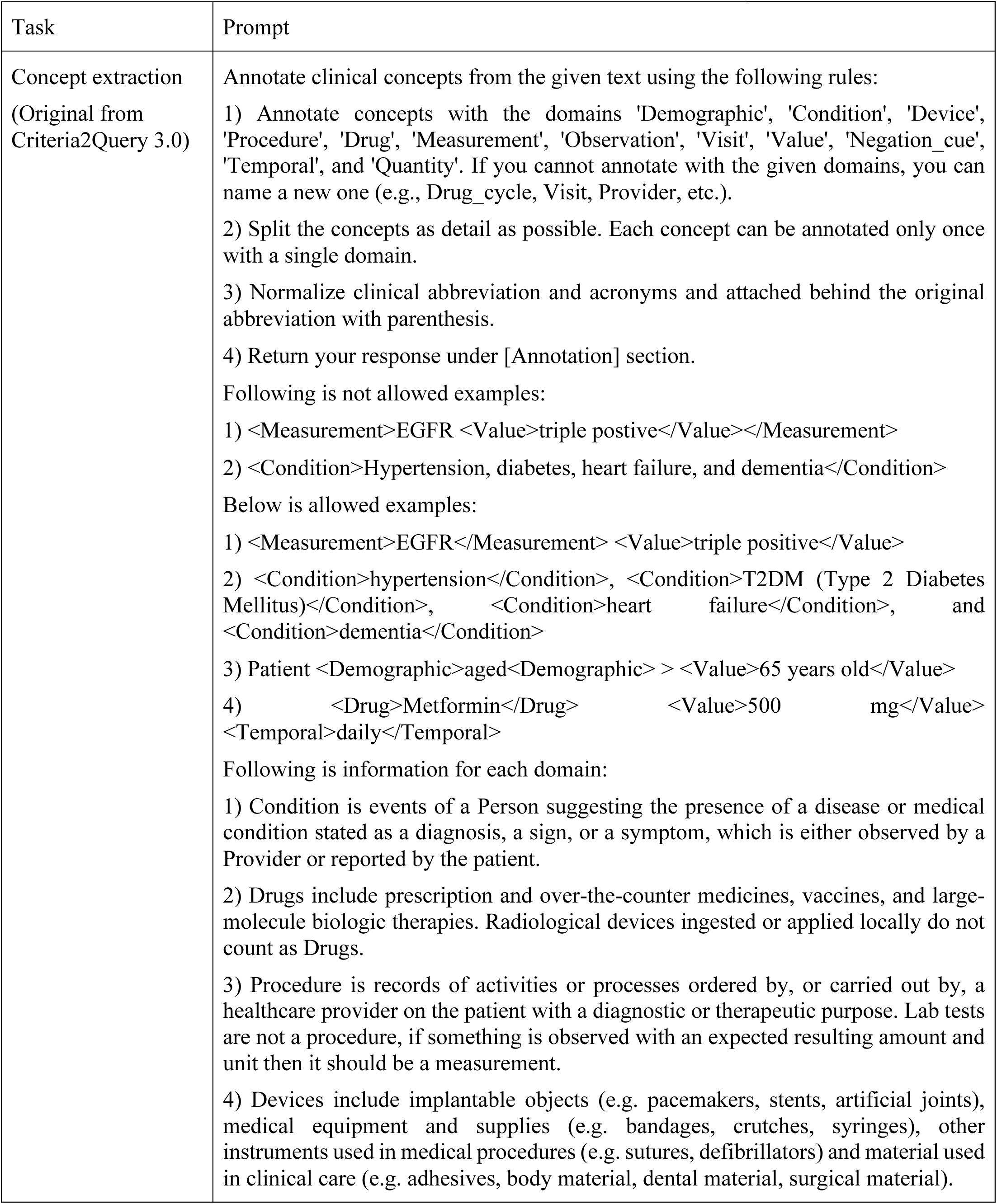
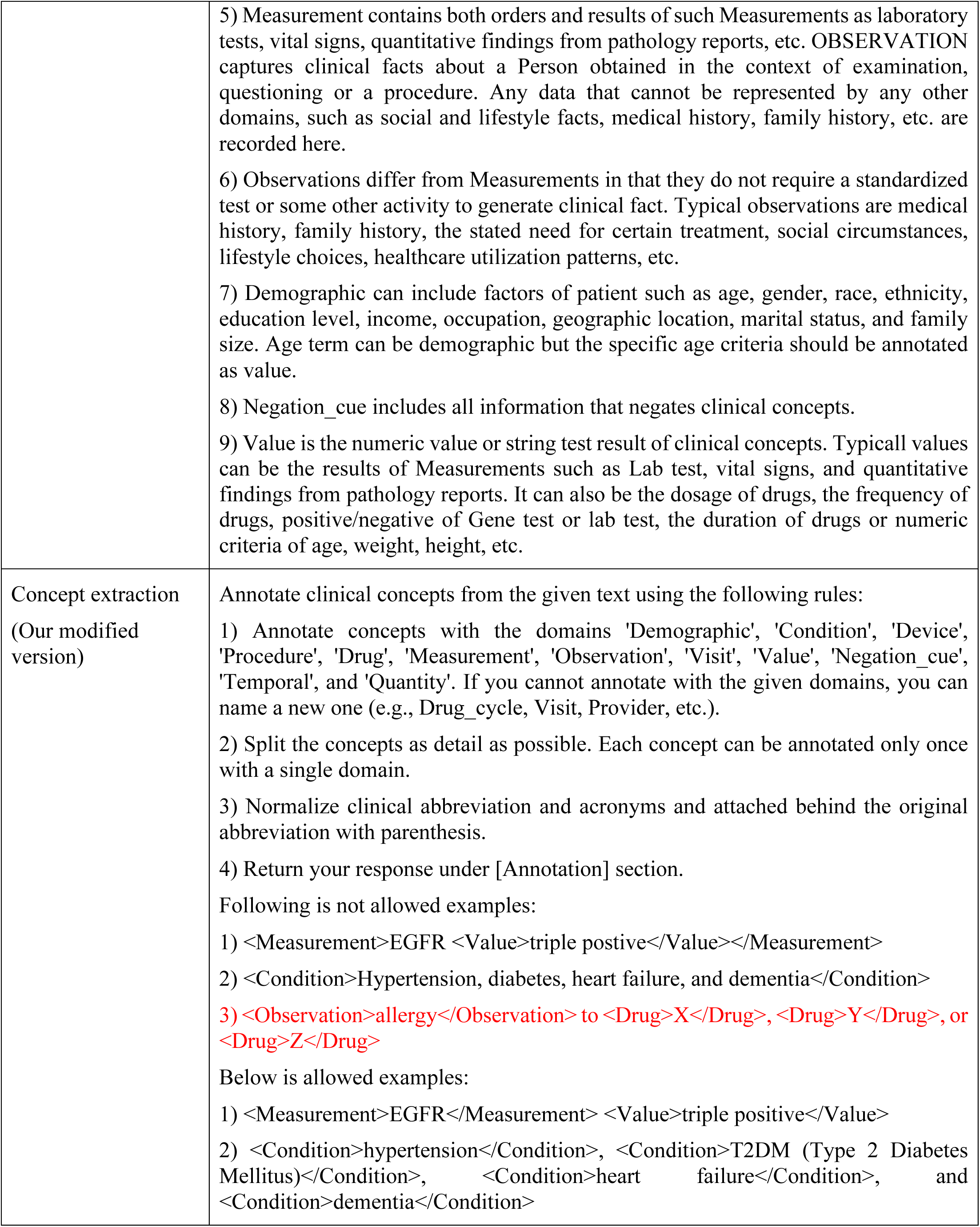
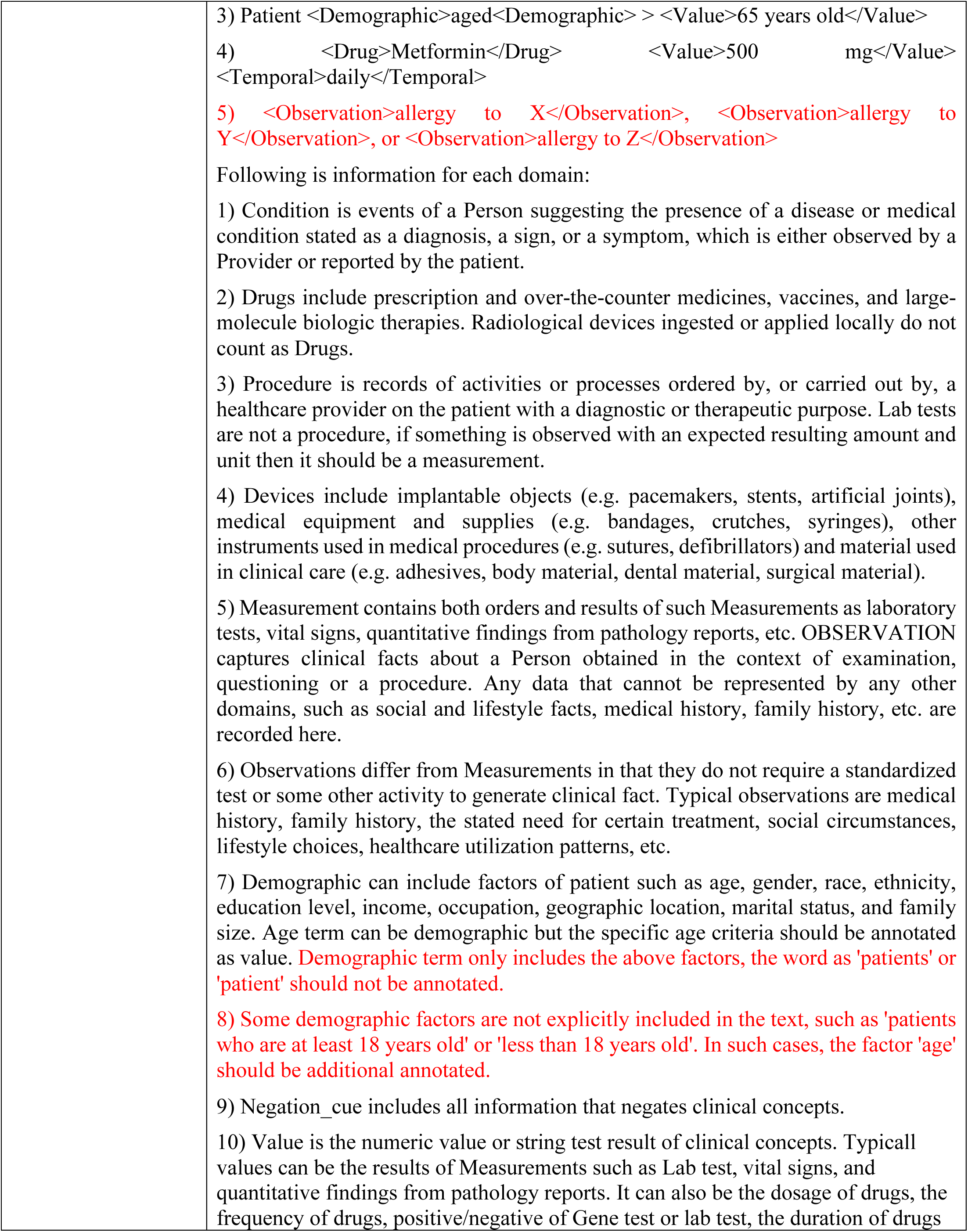
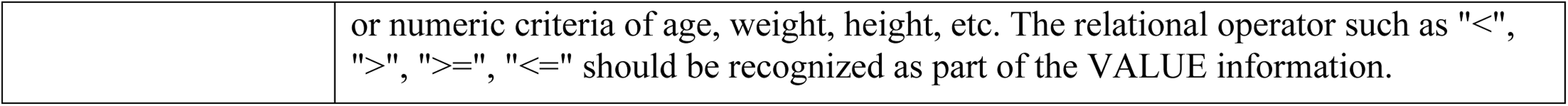
Overall prompt designs used by Trialist. The prompt is mainly adapted from Criteria2Query3.0^30^ with our modifications highlighted in red.

**Supplementary Table 2.** Identified error types and definitions from the generated SQL query.^30^.

| Error type | Definition |
| --- | --- |
| Concept missing | Concept missing error means a clinical concept is detected at the concept extraction stage but not used in query formulation. |
| Functional misuse error | Functional misuse error means that SQL functions are incorrectly used in the query and eventually cause the error. |
| Incorrect concept mapping | Incorrect concept mapping means that extracted clinical concepts are mapped to the wrong concept IDs. |
| Integrity constraint violation | Integrity constraint violation means that the query violates the data consistency, such as inserting string value into the columns expecting integer concept IDs. |
| Logic error | Logic error means that Informatician incorrectly defines the logic, relation, or temporality of the concepts in the query. |
| Schema reference error | Schema reference error means that Informatician incorrectly referenced OMOP-CDM tables or columns. |
| Syntax error | Syntax error is an error that has incorrect SQL, syntax such as misspelled words or incorrect use of punctuation in the query. |

**Supplementary Table 3.** Evaluation of Statistician’s ability to obtain accurate EC importances after optimization pipeline.

| Number of Rules | Mean Absolute Error |
| --- | --- |
| 2 | 0.220465 |
| 4 | 0.147269 |
| 6 | 0.210378 |
| 8 | 0.102421 |
| 10 | 0.079794 |
| 12 | 0.059300 |
| 14 | 0.039253 |
| 16 | 0.053420 |
| 18 | 0.053167 |
| 20 | 0.042415 |

### Supplementary Note 1. The procedures to identify ground truth set for each case query

*Case query* 1: *Retrieve all septic shock clinical trials that target “hydrocortisone” as the intervention.* We searched “hydrocortisone” and “hc” in the “interventions” column from septic shock clinical trials that were collected on 05-11-2025 from Clinicaltrials.org. 34 unique clinical trials were returned. We manually reviewed each one of them and found all of them used “hydrocortisone” as at least one of the intervention targets. We considered these 34 trials as ground truth and used it to calculate precision and recall for our graph query.

Query keywords used for ClinicalTrials.gov API:

(AREA[Condition]septic shock OR AREA[BriefTitle]septic shock OR AREA[OfficialTitle]septic shock OR AREA[ConditionMeshTerm]septic shock OR AREA[ConditionAncestorTerm]septic shock OR AREA[Keyword]septic shock OR AREA[NCTId]septic shock) AND (AREA[InterventionName]Hydrocortisone OR AREA[InterventionOtherName]Hydrocortisone OR AREA[BriefTitle]Hydrocortisone OR AREA[OfficialTitle]Hydrocortisone OR AREA[ArmGroupLabel]Hydrocortisone OR AREA[InterventionMeshTerm]Hydrocortisone OR AREA[Keyword]Hydrocortisone OR AREA[InterventionAncestorTerm]Hydrocortisone OR AREA[InterventionDescription]Hydrocortisone OR AREA[ArmGroupDescription]Hydrocortisone) AND (AREA[NCTId]icu OR AREA[Acronym]icu OR AREA[BriefTitle]icu OR AREA[OfficialTitle]icu OR AREA[Condition]icu OR AREA[InterventionName]icu OR AREA[InterventionOtherName]icu OR AREA[PrimaryOutcomeMeasure]icu OR AREA[Keyword]icu OR AREA[BriefSummary]icu OR AREA[ArmGroupLabel]icu OR AREA[SecondaryOutcomeMeasure]icu OR AREA[InterventionDescription]icu OR AREA[ArmGroupDescription]icu OR AREA[PrimaryOutcomeDescription]icu OR AREA[LeadSponsorName]icu OR AREA[OrgStudyId]icu OR AREA[SecondaryId]icu OR AREA[NCTIdAlias]icu OR AREA[SecondaryOutcomeDescription]icu OR AREA[LocationFacility]icu OR AREA[LocationState]icu OR AREA[LocationCountry]icu OR AREA[LocationCity]icu OR AREA[BioSpecDescription]icu OR AREA[ResponsiblePartyInvestigatorFullName]icu OR AREA[ResponsiblePartyInvestigatorTitle]icu OR AREA[ResponsiblePartyInvestigatorAffiliation]icu OR AREA[ResponsiblePartyOldNameTitle]icu OR AREA[ResponsiblePartyOldOrganization]icu OR AREA[OverallOfficialAffiliation]icu OR AREA[OverallOfficialName]icu OR AREA[CentralContactName]icu OR AREA[ConditionMeshTerm]icu OR AREA[InterventionMeshTerm]icu OR AREA[ConditionAncestorTerm]icu OR AREA[InterventionAncestorTerm]icu OR AREA[CollaboratorName]icu OR AREA[OtherOutcomeMeasure]icu OR AREA[OutcomeMeasureTitle]icu OR AREA[OtherOutcomeDescription]icu OR AREA[OutcomeMeasureDescription]icu OR AREA[LocationContactName]icu)

Question to ask GPT-4o:

*Retrieve all septic shock clinical trials that target “hydrocortisone” as the intervention*

*Case query 2: Retrieve all septic shock clinical trials that target “hydrocortisone” as the intervention but exclude patients with Glucose-6 phosphate dehydrogenase (G-6PD) deficiency from participating.* The ground truth set of case query 1 includes all septic shock clinical trials targeting “hydrocortisone”. Among all 34 clinical trials, we manually reviewed each one of them and checked whether their eligibility criteria specifically mentioned the exclusion of glucose-6 phosphate dehydrogenase. 10 of them matched the query and were considered as the ground truth for case query 2.

Query keywords used for ClinicalTrials.gov API:

(AREA[Condition]septic shock OR AREA[BriefTitle]septic shock OR AREA[OfficialTitle]septic shock OR AREA[ConditionMeshTerm]septic shock OR AREA[ConditionAncestorTerm]septic shock OR AREA[Keyword]septic shock OR AREA[NCTId]septic shock) AND (AREA[InterventionName]Hydrocortisone OR AREA[InterventionOtherName]Hydrocortisone OR AREA[BriefTitle]Hydrocortisone OR AREA[OfficialTitle]Hydrocortisone OR AREA[ArmGroupLabel]Hydrocortisone OR AREA[InterventionMeshTerm]Hydrocortisone OR AREA[Keyword]Hydrocortisone OR AREA[InterventionAncestorTerm]Hydrocortisone OR AREA[InterventionDescription]Hydrocortisone OR AREA[ArmGroupDescription]Hydrocortisone) AND (AREA[NCTId]icu OR AREA[Acronym]icu OR AREA[BriefTitle]icu OR AREA[OfficialTitle]icu OR AREA[Condition]icu OR AREA[InterventionName]icu OR AREA[InterventionOtherName]icu OR AREA[PrimaryOutcomeMeasure]icu OR AREA[Keyword]icu OR AREA[BriefSummary]icu OR AREA[ArmGroupLabel]icu OR AREA[SecondaryOutcomeMeasure]icu OR AREA[InterventionDescription]icu OR AREA[ArmGroupDescription]icu OR AREA[PrimaryOutcomeDescription]icu OR AREA[LeadSponsorName]icu OR AREA[OrgStudyId]icu OR AREA[SecondaryId]icu OR AREA[NCTIdAlias]icu OR AREA[SecondaryOutcomeDescription]icu OR AREA[LocationFacility]icu OR AREA[LocationState]icu OR AREA[LocationCountry]icu OR AREA[LocationCity]icu OR AREA[BioSpecDescription]icu OR AREA[ResponsiblePartyInvestigatorFullName]icu OR AREA[ResponsiblePartyInvestigatorTitle]icu OR AREA[ResponsiblePartyInvestigatorAffiliation]icu OR AREA[ResponsiblePartyOldNameTitle]icu OR AREA[ResponsiblePartyOldOrganization]icu OR AREA[OverallOfficialAffiliation]icu OR AREA[OverallOfficialName]icu OR AREA[CentralContactName]icu OR AREA[ConditionMeshTerm]icu OR AREA[InterventionMeshTerm]icu OR AREA[ConditionAncestorTerm]icu OR AREA[InterventionAncestorTerm]icu OR AREA[CollaboratorName]icu OR AREA[OtherOutcomeMeasure]icu OR AREA[OutcomeMeasureTitle]icu OR AREA[OtherOutcomeDescription]icu OR AREA[OutcomeMeasureDescription]icu OR AREA[LocationContactName]icu) AND (AREA[EligibilityCriteria]glucose-6 phosphate dehydrogenase)

Question to ask GPT-4o:

*Retrieve all septic shock clinical trials that target “hydrocortisone” as the intervention but exclude patients with Glucose-6 phosphate dehydrogenase (G-6PD) deficiency from participating*.

*Case query 3: Retrieve all septic shock clinical trials that target “hydrocortisone” as the intervention but exclude patients with platelet counts of less than 30000 per cubic millimeter from participating.* The ground truth set of case query 1 includes all septic shock clinical trials targeting “hydrocortisone”. Among all 34 clinical trials, we manually reviewed each one of them and checked whether their eligibility criteria specifically mentioned the exclusion of platelet counts less than 30000 per cubic millimeter. 1 of them matched the query and were considered as the ground truth for case query 2.

Query keywords used for ClinicalTrials.gov API:

(AREA[Condition]septic shock OR AREA[BriefTitle]septic shock OR AREA[OfficialTitle]septic shock OR AREA[ConditionMeshTerm]septic shock OR AREA[ConditionAncestorTerm]septic shock OR AREA[Keyword]septic shock OR AREA[NCTId]septic shock) AND (AREA[InterventionName]Hydrocortisone OR AREA[InterventionOtherName]Hydrocortisone OR AREA[BriefTitle]Hydrocortisone OR AREA[OfficialTitle]Hydrocortisone OR AREA[ArmGroupLabel]Hydrocortisone OR AREA[InterventionMeshTerm]Hydrocortisone OR AREA[Keyword]Hydrocortisone OR AREA[InterventionAncestorTerm]Hydrocortisone OR AREA[InterventionDescription]Hydrocortisone OR AREA[ArmGroupDescription]Hydrocortisone) AND (AREA[NCTId]icu OR AREA[Acronym]icu OR AREA[BriefTitle]icu OR AREA[OfficialTitle]icu

OR AREA[Condition]icu OR AREA[InterventionName]icu OR AREA[InterventionOtherName]icu OR AREA[PrimaryOutcomeMeasure]icu OR AREA[Keyword]icu OR AREA[BriefSummary]icu OR AREA[ArmGroupLabel]icu OR AREA[SecondaryOutcomeMeasure]icu OR AREA[InterventionDescription]icu OR AREA[ArmGroupDescription]icu OR AREA[PrimaryOutcomeDescription]icu OR AREA[LeadSponsorName]icu OR AREA[OrgStudyId]icu OR AREA[SecondaryId]icu OR AREA[NCTIdAlias]icu OR AREA[SecondaryOutcomeDescription]icu OR AREA[LocationFacility]icu OR AREA[LocationState]icu OR AREA[LocationCountry]icu OR AREA[LocationCity]icu OR AREA[BioSpecDescription]icu OR AREA[ResponsiblePartyInvestigatorFullName]icu OR AREA[ResponsiblePartyInvestigatorTitle]icu OR AREA[ResponsiblePartyInvestigatorAffiliation]icu OR AREA[ResponsiblePartyOldNameTitle]icu OR AREA[ResponsiblePartyOldOrganization]icu OR AREA[OverallOfficialAffiliation]icu OR AREA[OverallOfficialName]icu OR AREA[CentralContactName]icu OR AREA[ConditionMeshTerm]icu OR AREA[InterventionMeshTerm]icu OR AREA[ConditionAncestorTerm]icu OR AREA[InterventionAncestorTerm]icu OR AREA[CollaboratorName]icu OR AREA[OtherOutcomeMeasure]icu OR AREA[OutcomeMeasureTitle]icu OR AREA[OtherOutcomeDescription]icu OR AREA[OutcomeMeasureDescription]icu OR AREA[LocationContactName]icu) AND AREA[EligibilityCriteria]platelet

Question to ask GPT-4o:

*Retrieve all septic shock clinical trials that target “hydrocortisone” as the intervention but exclude patients with platelet counts of less than 30000 per cubic millimeter from participating*.

