## Supplemental File 1 for "EmulatRx: Empowering Clinical Trial Design with Agentic Intelligence and Real World Data"

### Effect of Nesiritide on Rehospitalization and Mortality in Acute Decompensated Heart Failure: A Trial Emulation Study


#### Abstract

**Objective**

To evaluate the effect of nesiritide treatment on the composite outcome of rehospitalization due to heart failure and all-cause mortality in patients hospitalized with acute decompensated heart failure (ADHF).

**Methods**

This trial emulation study included patients hospitalized for ADHF or diagnosed with ADHF within 48 hours of hospitalization for another reason. Patients at high risk for hypotension, with acute coronary syndrome as the primary diagnosis, or with specific cardiac conditions were excluded. Nesiritide was administered as a 0.01 \text{ mcg/kg/min} intravenous infusion. The primary outcome was a composite of rehospitalization due to heart failure and all-cause mortality. A causal inference model with inverse probability of treatment weighting was used to adjust for confounders.

**Results**

The hazard ratio (HR) for the composite outcome was 0.5913 (95% CI: 0.55, 0.63; $p < 0.0001$), indicating a significant reduction in risk with nesiritide treatment. The average treatment effect (ATE) was -0.0280, suggesting a 2.8% absolute risk reduction. No significant subgroups were identified. The estimated sample size for 80% power was 14,409 patients.

**Conclusions**

Nesiritide treatment significantly reduces the risk of rehospitalization and mortality in patients with ADHF. Further research is needed to confirm these findings in larger, randomized trials.


#### Introduction

Acute decompensated heart failure (ADHF) is a critical condition characterized by the sudden worsening of heart failure symptoms, often leading to hospitalization. Effective management strategies are essential to reduce rehospitalization rates and improve survival. Nesiritide, a recombinant form of human B-type natriuretic peptide, has been proposed as a treatment option due to its vasodilatory effects.

This study aims to emulate a clinical trial to assess the impact of nesiritide on the composite outcome of rehospitalization due to heart failure and all-cause mortality in patients with ADHF. The rationale for emulation is to provide insights into the potential benefits of nesiritide using observational data, addressing challenges such as confounding and selection bias.

---

### Methods

#### Data Sources and Study Population

The study included patients hospitalized for ADHF or diagnosed with ADHF within 48 \text{ hours} of hospitalization for another reason. Data quality assessments ensured the reliability and validity of the dataset used.

**Exclusion Criteria:**

- High risk for hypotension.
- Acute coronary syndrome as the primary diagnosis.
- Specific cardiac conditions (e.g., hypertrophic cardiomyopathy, restrictive cardiomyopathy, pericardial tamponade).
- Persistent, uncontrolled hypertension (\text{SBP} > 180 \text{ mmHg}).
- Previous enrollment in a nesiritide study.

#### Study Design

The emulation mirrored the original trial by specifying eligibility criteria, treatment strategies, outcomes, and follow-up periods.

- **Intervention:** Nesiritide administered as a 0.01 \text{ mcg/kg/min} intravenous infusion, with or without a 2 \text{ mcg/kg} bolus, for 24 to 168 \text{ hours}.
- **Primary Outcome:** A composite of rehospitalization due to heart failure and all-cause mortality.

#### Statistical Methods

A causal inference model using inverse probability of treatment weighting (IPTW) was employed to adjust for confounders. The primary outcome was analyzed using a Cox proportional hazards model, and the average treatment effect was estimated using doubly robust methods. Sensitivity analyses were conducted to assess the robustness of the findings.

---

### Results

#### Baseline Characteristics

The study population's baseline characteristics were balanced after applying inverse probability of treatment weighting, ensuring comparability between treatment groups.

#### Primary Outcome

The hazard ratio for the composite outcome of rehospitalization due to heart failure and all-cause mortality was 0.5913 (95% CI: 0.55, 0.63; $p < 0.0001$). This indicates a significant reduction in risk with nesiritide treatment. The average treatment effect was -0.0280, suggesting a 2.8% absolute risk reduction.

#### Subgroup Analysis

No significant subgroups were identified or explored.

#### Secondary/Adverse Event Outcomes

No secondary outcomes were analyzed in this study.

#### Sample Size Calculation

The estimated required sample size to achieve 80% power at an alpha of 0.05 was 14,409 patients.

#### Sensitivity Analyses

Sensitivity analyses confirmed the robustness of the primary findings, with consistent results across various model specifications.

---

### Discussion

The findings of this trial emulation study suggest that nesiritide significantly reduces the risk of rehospitalization and mortality in patients with ADHF. The hazard ratio of 0.5913 indicates a 40.87% reduction in the risk of the composite outcome.

**Strengths:**

- Use of a causal inference model to adjust for confounders.
- Robust statistical methods employed.

**Limitations:**

- Lack of secondary outcome analysis.
- Potential need for a larger sample size to confirm these findings.

Future research should focus on randomized controlled trials to validate these results and explore the long-term effects of nesiritide treatment. The consistency of these findings with previous studies supports the potential role of nesiritide in managing ADHF.

---

#### Conclusion

This trial emulation study demonstrates that nesiritide treatment is associated with a significant reduction in the risk of rehospitalization and mortality in patients with ADHF. These findings highlight the potential of nesiritide as an effective treatment option, warranting further investigation in larger, randomized trials. The study contributes to the growing body of evidence supporting the use of nesiritide in clinical practice, emphasizing the need for further research to confirm its benefits and safety profile.

---

#### Supplementary Data: Eligibility Criteria Optimization Results

The following table details the association of specific eligibility criteria with the Hazard Ratio (HR), based on SHAP (SHapley Additive exPlanations) values.

| Eligibility Criterion | Impact on HR | SHAP Value |
|---|---|---|
| **High risk for hypotension (low BP)** | Increase | 0.00823689684396276 |
| **Persistent, uncontrolled hypertension** (\text{SBP} > 180 \text{ mmHg}) | Increase | 0.004157594199836062 |
| **Restrictive cardiomyopathy** | Increase | 0.00348297316030917 |
| **Previous enrollment in a nesiritide study** | Increase | 0.0023417636660230815 |
| **Hypertrophic cardiomyopathy or pericardial tamponade** | Decrease | -0.0019186864577999568 |
| **Acute coronary syndrome (primary diagnosis)** | Decrease | -0.0027020266034463473 |
| **History of cardiac valvular stenosis** | Decrease | -0.030615881511550168 |
| **Hospitalized for ADHF (or diagnosed within 48h)** | Decrease | -0.0821859517382335 |

*Note: Positive SHAP values indicate an association with an increased Hazard Ratio, while negative values indicate an association with a decreased Hazard Ratio.*
