## Supplemental File 3 for "EmulatRx: Empowering Clinical Trial Design with Agentic Intelligence and Real World Data"

### Effectiveness of Hydrocortisone Sodium Succinate in Reducing 28-Day Mortality in ICU Patients with Septic Shock: A Trial Emulation Study


#### Abstract

###### Objective

To evaluate the effect of hydrocortisone sodium succinate on 28-day mortality in patients with septic shock admitted to the ICU.

###### Methods

This trial emulation study included patients aged 18 years or older admitted to the ICU with septic shock within 24 hours, as defined by SEPSIS-3 criteria. The intervention group received hydrocortisone sodium succinate at a dose of 50 mg every 6 hours for 7 days or until ICU discharge. The primary outcome was 28-day mortality. A causal inference model was used to estimate the treatment effect, adjusting for potential confounders. Inverse Probability of Treatment Weighting (IPTW) and Cox Proportional Hazards models were employed.

###### Results

The hazard ratio (HR) for 28-day mortality was 1.2983 (95% CI: 1.0700, 1.5753; $p = 0.0082$), indicating a 29.83% increased risk of mortality in the treatment group. The average treatment effect (ATE) was 0.115 (95% CI: 0.043, 0.181). No significant subgroups were identified. The estimated sample size required for 80% power was 2182 patients.

###### Conclusions

Hydrocortisone sodium succinate was associated with an increased risk of 28-day mortality in ICU patients with septic shock. Further research is needed to explore these findings and assess the potential for unmeasured confounding.


---

#### Introduction

Septic shock is a critical condition characterized by severe infection leading to systemic inflammation and organ dysfunction. Despite advances in critical care, mortality rates remain high. Corticosteroids, such as hydrocortisone, have been proposed as adjunctive therapy to modulate the inflammatory response.

This study aims to emulate a clinical trial to assess the effectiveness of hydrocortisone sodium succinate in reducing 28-day mortality among ICU patients with septic shock. The rationale for this emulation is to provide insights into the potential benefits and risks of corticosteroid use in this population, given the mixed results from previous trials.

### Methods

#### Data Sources and Study Population

The study included patients aged 18 years or older admitted to the ICU with septic shock within 24 hours, as defined by SEPSIS-3 criteria.

**Exclusion Criteria:**

- Pregnancy
- Known G6PD deficiency
- Acute stroke
- Acute coronary syndrome
- Active gastrointestinal bleed
- Burn
- Trauma
- Vasopressor use prior to randomization for more than 24 hours

Data quality assessments ensured the reliability and validity of the dataset used.

#### Intervention and Comparator

- **Intervention:** Hydrocortisone sodium succinate at a dose of 50 mg every 6 hours for 7 days or until ICU discharge.
- **Comparator:** Standard care without hydrocortisone.

#### Statistical Methods

A causal inference model was used to estimate the treatment effect on 28-day mortality, adjusting for potential confounders. Inverse Probability of Treatment Weighting was applied to balance covariates. The Cox Proportional Hazards model provided hazard ratios, and Doubly Robust Estimation was used to confirm findings. Sensitivity analyses included E-Value calculations to assess the impact of unmeasured confounding.

### Results

#### Baseline Characteristics

The study population was well-characterized, with balanced covariates achieved through weighting techniques.

#### Primary Outcome

The hazard ratio for 28-day mortality was 1.2983 (95% CI: 1.0700, 1.5753; $p = 0.0082$). This indicates a 29.83% increased risk of mortality in the treatment group. The average treatment effect was 0.115 (95% CI: 0.043, 0.181).

##### Subgroup Analysis

No significant subgroups were identified.

##### Secondary/Adverse Event Outcomes

No secondary outcomes were analyzed.

##### Sample Size Calculation

The estimated sample size required to achieve 80% power at an alpha of 0.05 was 2182 patients.

##### E-Value Calculation

The minimum strength of association that an unmeasured confounder would need to have with both the exposure and the outcome to completely eliminate the observed effect was 1.9341.

---

#### Discussion

The study found that hydrocortisone sodium succinate was associated with an increased risk of 28-day mortality in ICU patients with septic shock. The hazard ratio of 1.2983 suggests a 29.83% increased risk of mortality, which is statistically significant.

**Strengths:**

- Use of a causal inference model.
- Robust statistical methods to adjust for confounders.

**Limitations:**

- Presence of unbalanced covariates, such as absolute eosinophils.
- Potential for unmeasured confounding.
- The required sample size for adequate power was not met, which may affect the reliability of the findings.

Future research should focus on larger sample sizes and explore potential confounders. Comparisons with original trials indicate consistency in the direction of the effect, though the magnitude differs, highlighting the need for further investigation.

---

#### Conclusion

This trial emulation study suggests that hydrocortisone sodium succinate may increase the risk of 28-day mortality in ICU patients with septic shock. These findings highlight the need for caution in the use of corticosteroids in this population and underscore the importance of further research to confirm these results and explore underlying mechanisms. The study contributes to the ongoing debate on corticosteroid use in septic shock and emphasizes the need for well-powered trials to validate these findings.

---

#### Supplementary Data: Eligibility Criteria Optimization Results

The following table details the association of specific eligibility criteria with the Hazard Ratio (HR), based on SHAP (SHapley Additive exPlanations) values.

| Eligibility Criterion | Impact on HR | SHAP Value |
|---|---|---|
| **Septic shock (SEPSIS-3)** | Increase | 0.3720194966219966 |
| **Trauma** | Increase | 0.10695848156712562 |
| **Acute coronary syndrome** | Increase | 0.016166480463808246 |
| **Vasopressor use (>24h pre-randomization)** | Increase | 0.01579768597575906 |
| **Pregnant (negative serum HCG)** | Increase | 0.0022538580293985843 |
| **Known G6PD deficiency** | Increase | 0.0021499878824924492 |
| **Age** >= 18 years | Neutral | 0.0 |
| **Burn** | Decrease | -0.00022287649487537343 |
| **Active gastrointestinal bleed** | Decrease | -0.001470872363434779 |
| **Acute stroke** | Decrease | -0.003052664661016897 |

*Note: Positive SHAP values indicate an association with an increased Hazard Ratio, while negative values indicate an association with a decreased Hazard Ratio.*
